# EXaCT-2: An augmented and customizable oncology-focused whole exome sequencing platform

**DOI:** 10.1101/2024.12.05.24318515

**Authors:** Peter Waltman, Pooja Chandra, Ken W Eng, David C Wilkes, Hyeon Park, Carlos Pabon, Princesca Delpe, Bhavneet Bhinder, Jyothi Manohar, Troy Kane, Evan Fernandez, Kathryn Gorski, Noah Greco, Manuele Simi, Jeffrey M Tang, Pantelis Zisimopoulos, Abigail King, Majd Al Assaad, Theresa Teneyck, Douglas Roberts, Jorge Monge, Francesca Demichelis, Wayne Tam, Madhu M Ouseph, Alexandros Sigaras, Himisha Beltran, Hannah Rennert, Neal Lindeman, Wei Song, James Solomon, Juan Miguel Mosquera, Rob Kim, Jeffrey Catalano, Duane C Hassane, Michael Sigouros, Olivier Elemento, Alicia Alonso, Andrea Sboner

**Author notes:** Alicia Alonso and Andrea Sboner jointly supervised this work. **Corresponding Author** Andrea Sboner, PhD, Department of Pathology and Laboratory Medicine, Englander Institute of Precision Medicine, Institute for Computational Biomedicine, Meyer Cancer Center, Weill Cornell Medicine, New York, NY, Mastodon handle: @.

## Abstract

With the rapid advances in cancer research, the list of variants and genes that drive human diseases is constantly expanding. Moreover, the FDA has approved more cancer therapies that incorporate a broader set of genomic features than simple gene variants such as Tumor Mutation Burden (TMB), microsatellite instability status (MSI), and fusion events in gene families such as the NTRK receptors. These features currently require multiple testing methods (IHC/FISH/etc.). With the cost of NGS testing dropping, it is now possible to envision an NGS assay capable of reliably detecting these features without the need for additional testing. The EIPM multidisciplinary team has developed EXaCT-2: a whole exome sequencing (WES) assay that gives the coverage of a targeted assay on cancer genes and the breadth to detect copy number events, cancer-related fusions, and viruses, which can facilitate diagnostic and therapeutic decisions for cancer patients.

We evaluated EXaCT-2 on 250 matched tumor/normal pairs and compared its performance with orthogonally validated results. We show the assay achieves the expected coverage of critical cancer genes, provides a better characterization of somatic copy number alterations, detects common cancer rearrangements and viruses, and enables the accurate estimation of global molecular metrics, such as tumor mutational burden and microsatellite instability. Further, we demonstrate the sensitivity of the assay to identify sub-clonal mutations that standard whole-exome assays are incapable of detecting, including the presence of KRAS mutations in samples previously believed to only contain wild-type KRAS, as well aging-related, somatic mosaicism in a phenotypically benign sample that is proximal to endometrial cancer.

## Introduction

Advances in our understanding of the molecular basis of tumor development and progression have led to the identification of many alterations in the cancer genome, resulting in the development of targeted therapeutics^1–4^. The list of genes that drive the disease continues to expand, as does the number of potential treatment options available to patients, bringing us closer to fulfilling the promise of precision medicine: “administering the right drug to the right patient at the right time”. Indeed, response in phase I clinical trials is higher when treatment is based on biomarker eligibility^5–10^. Next-generation sequencing (NGS) provides a comprehensive evaluation of genomic alterations and inform diagnosis, prognosis, and therapeutic avenues and it is routinely available at many healthcare facilities and molecular laboratories. However, most oncology tests are based on a limited set of genes and variants (targeted panels), which leads to the following problems: 1) they need to be updated when new cancer variants are discovered. This a non-trivial issue as much of our knowledge of the disease is biased by using datasets enriched with people of European descent ^11–13^. As large-scale projects such as ICGC-ARGO, H3Africa, AllofUs^14–17^ work to address this bias, the field will need to quickly consider new additional diagnostically or therapeutically relevant cancer genes; 2) they tend to overestimate important features such as tumor mutational burden (TMB) due to the limited genomic territory they interrogate^18^. As new immunotherapy treatments become more available, accurate TMB estimation is important as an indicator of responsiveness to immunotherapy^19–22^; 3) they are limited in evaluating mutational signatures, a genomic feature increasingly relevant to the understanding of the underlying mechanisms leading to cancer development as well as the results of therapeutic intervention^23^; 4) impaired evaluation of somatic copy number alterations (SCNAs) due to limited information from broader genomic regions. Determining a single gene copy number status is possible but characterizing it as ‘focal’ (localized) or ‘broad’ (large areas or entire chromosomal arm) is difficult, with different disease implications^24–27^; 5) identification of gene rearrangements is limited, unless the associated breakpoints is targeted by the assay. As gene rearrangements are increasingly important biomarkers^28–30^, their omission may prove deleterious for many patients. Currently, they are identified as gene fusions via RNA profiling, hence requiring a different workflow in the lab, with potential delays in returning results^31, 32^.

Using broader NGS assays, such as WES and Whole Genome Sequencing (WGS) can overcome some of these limitations^33^. However, these assays are unable to achieve the depth of coverage that targeted panels can provide^33, 34^. Lower coverage can impair detection of important events in tumors with high normal cell contamination or subclonal events.

We here describe EXaCT-2, a new sequencing assay that combines the breadth of a WES assay with features of WGS and the depth of targeted panels (see Figure 1) that has already demonstrated its utility by one recent study^36^. Designed with cancer genomics in mind, EXaCT-2 aims for a coverage of ~400x for nearly 1,400 cancer genes, comparable to that of targeted panels, while still providing an average 100x coverage for the remainder of the exome (Table S1). Additionally, 32,700 probes on common SNPs, spread evenly across the genome, enable accurate estimation of both tumor purity and the boundaries of SCNAs. Furthermore, selected intronic areas are included to capture common rearrangement breakpoints, such as BRAF, and ALK. Similarly, immunoglobulin regions are almost entirely covered. Finally, EXaCT-2 can interrogate the presence of four common viruses involved in hematological cancers. In summary, we here present an assay that can interrogate multiple oncogenic events in one single assay, thus reducing the need for serial testing.

**Figure 1:**
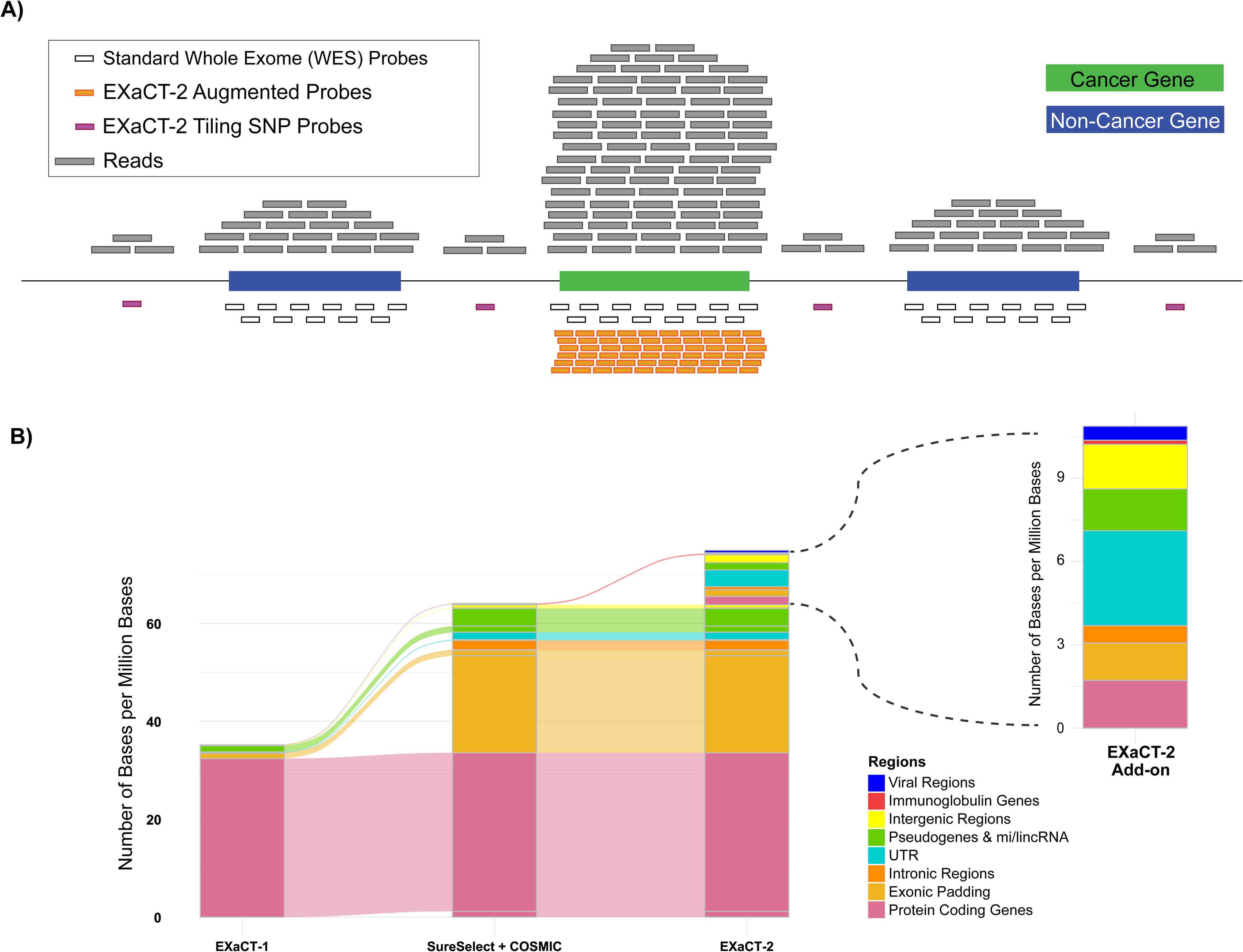
EXaCT-2 Assay Design. A) Schematic of the probe design strategy. B) Representation of the bases covered by EXaCT-2. The plot shows comparison with the whole exome assays EXaCT-1 and the standard SureSelect + Cosmic. The inset shows EXaCT-2 additional bases in specific regions such as intergenic and intronic areas, aiming to improve molecular profiling of specimens with enhanced resolution.

## Results

### Evaluation of novel features incorporated into EXaCT-2

The features relevant to cancer characterization as per FDA-approved drugs reflect a wide range of event types, from single nucleotide mutations to microsatellite instability, to complex rearrangements. They can be broadly categorized into the enhancement of both the Breadth of Coverage as well as the Depth of Coverage of the EXaCT-2 assay. Breadth of coverage improvements are achieved by the inclusion of probes for intronic regions (to allow for rearrangement detection), for intergenic regions (to improve on somatic copy number alteration detection), for comprehensive characterization of BCR rearrangements, and the inclusion of probes for known blood cancer-associated viruses. Depth of coverage improvements include modifications to boost the coverage of clinically relevant genes, such as EGFR and IDH2, as well as cancer genes difficult to assay due to GC-rich regions, such as CEBPA, KRAS, and PTEN. In the following sections, we use these categorizations to organize the discussion of our evaluation of these improvements.

### Breadth of Coverage

#### Improved genomic territory assayed by EXaCT-2

The EXaCT-2 assay we introduce here is a hybridization assay that builds upon the Agilent SureSelect + Cosmic assay (Santa Clara, CA) ^37^. To illustrate the novelty of EXaCT-2 we also compared it with EXaCT-1, a WES amplicon-based assay. Figure 1 illustrates the difference of the assays with respect to the coverage of the human genome (Table S2). The three assays remain relatively unchanged in terms of protein-coding regions (~21K genes – 35MB). Only a 3.6% increase in protein coding bases is targeted by EXaCT-2 and the SureSelect assays, compared to EXaCT-1. Due to the hybridization capture method, exonic padding regions, defined as the 50 non-coding bases flanking an exon, account for about 22MB of the assay compared to EXaCT-1.

As shown in Figure 1 - inset, the EXaCT-2 specific 10MB enhanced design includes: several intergenic targets for well-established non-coding SNPs to allow for accurate estimation of copy number boundaries resulting in an expansion of overlapping non-coding RNAs; probes for immunoglobulin regions that were previously missing – the inclusion of which allows for accurate estimation of clonal BCR rearrangements; probes for intronic hotspots of DNA breakpoints, such as the KIAA1549-BRAF; probes for 4 viral agents commonly involved in hematological or soft-tissue tumors, and probes for expanded coverage of the non-coding UTR regions, to improve the detection of mutations that impact mechanisms of post-transcriptional control.

#### Increased breadth of coverage improves resolution of somatic copy number alteration (SCNA) boundaries

As described above, we illustrate in Figure 2 the effect of the EXaCT-2 extended genomic territory on the assessment of the boundaries of somatic copy number alterations (SCNAs) by CNVKit (see Methods). In this example, EXaCT-1 and EXaCT-2 were applied to the same sample, with their respective analytical pipelines, identifying a somatic deletion involving the Desmocollin genes (DSC1, DSC2, DSC3). However, the deletion boundaries identified by the two pipelines were markedly different. The EXaCT-1 pipeline inferred a deletion boundary within the second exon of Cadherin 2 (CDH2; 18:25727685), while the EXaCT-2’s inclusion of intergenic SNPs allowed its pipeline to locate the boundary within the intergenic region (18:27646011) (Figure 2B).

**Figure 2:**
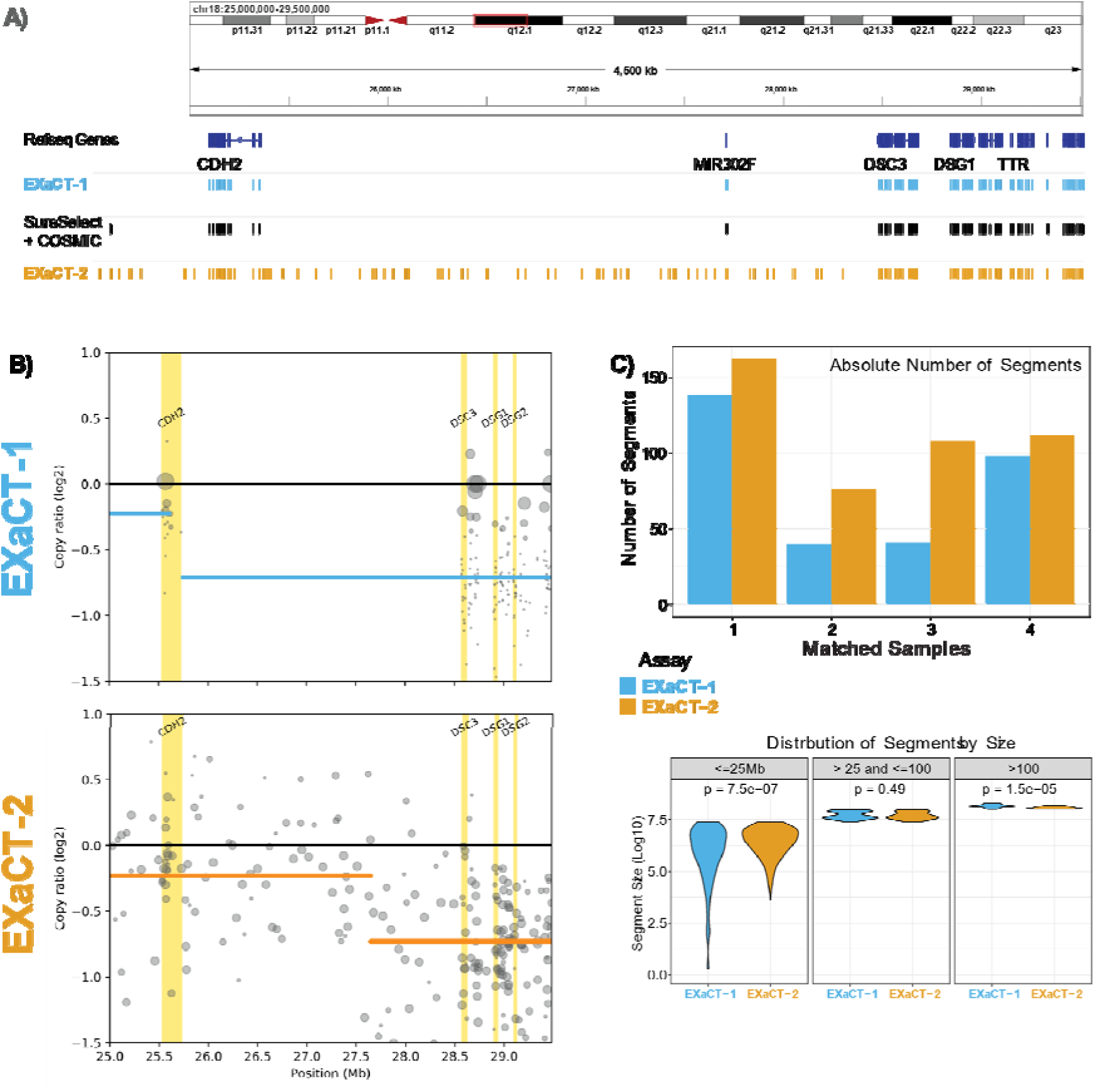
Somatic Copy Number Alteration (SCNAs) comparison between matching samples. **A)** Illustration of the same genomic region on chromosome 18 analyzed using EXaCT-1 and EXaCT-2 on the same sample, showing the corresponding genes and probe coverage across three different WES assays: EXaCT-1, SureSelect + COSMIC, and EXaCT-2. **B)** Segmented bin-level coverage profiles for EXaCT-1 (top) and EXaCT-2 (bottom). Note, the size of the data point is proportional to the weight of that point in the segmentation performed by CNVkit (see methods). **C – top)** Absolute Number of segments in EXaCT-1 and EXaCT-2 for the 4 matching samples. **C – bottom)** Comparison of segment sizes within the 3 categories of small, medium, and large, with statistical difference assessed by t-test.

Beyond this one example, a systematic comparison of 4 samples that were analyzed with both EXaCT-2 and EXaCT-1 was performed. This comparison demonstrated that while the segmentations that were generated from the two assays were highly concordant in terms of gene level alterations (Table S3), the inclusion of probes covering intergenic SNPs allows the segmentation from the EXaCT-2 assay to be much less under- and over-segmented than is possible with an exome-based assay like EXaCT-1 (Figures 2C, Figure S1-S2, Tables S4-S5).

#### Detection of rearrangements in common cancer driver genes

The EXaCT-2 assay evaluated 66 genes often involved in clinically-relevant gene fusions (Table S6) and covers a broad spectrum of cancer types (Figure S3).^38^ As this is a DNA-based assay, the assumptions are that: 1. genomic rearrangements are the basis for the generation of gene fusions, and 2. rearrangement breakpoints frequently occur within specific introns. With these two assumptions, we designed probes to cover introns that harbor some of the most common breakpoints (Table S7). Despite the limited scope of these non-coding regions, our evaluation indicated that their inclusion was remarkably effective (see Figure 3A for an example: KIAA1549-BRAF, a clinical marker in Pediatric Low-Grade Astrocytoma^28^). Using a set of ten positive control samples identified using traditional RNA-seq-based methods, the EXaCT-2 assay identified all 10 fusions (Tables S8 and S9).

**Figure 3:**
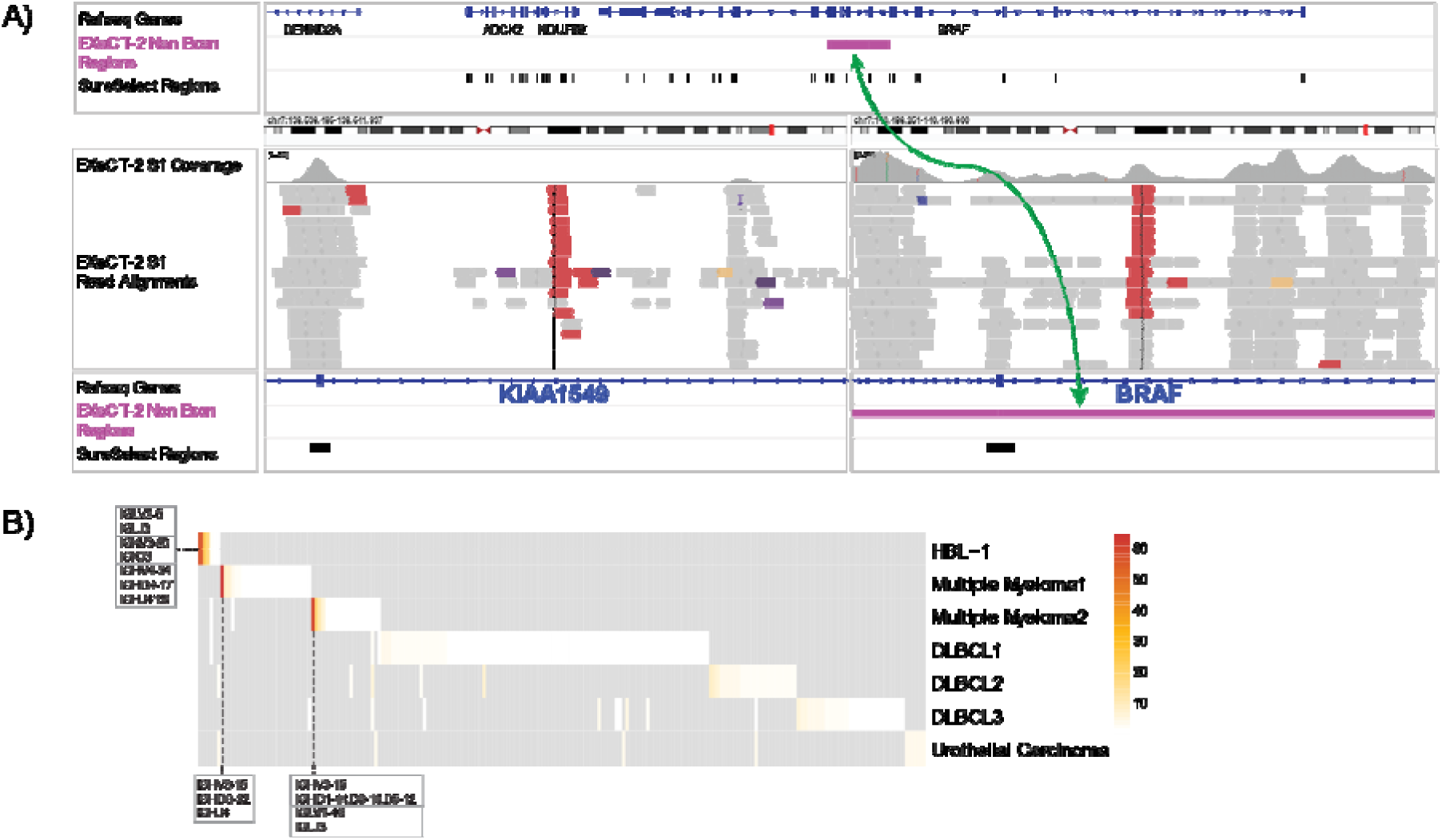
Types of rearrangements captured by EXaCT-2. **A) BRAF rearrangement.** In pink, a region known to be fusion hotspot for BRAF is captured by EXaCT-2. Below that is an IGV representation of BRAF-KIAA1549 rearrangement: Reads involved in the rearrangements are in red. Note, that the reads in the KIAA1549 locus align to an untargeted intronic region and are only recovered due the BRAF intronic region targeted by EXaCT-2. **B) BCR rearrangement.** Heatmap indicating the relative read frequency of each BCR clonal rearrangement within the corresponding sample. Gray areas represent a read frequency of 0, indicating the absence of that clonotype in the sample. Additionally, all the highlighted rearrangements in the box have a frequency of >= 15%.

#### Improved B-Cell Receptor (BCR) rearrangement detection

B cell development involves the coordinated rearrangement of Immunoglobulin (Ig) genes to produce functional antibodies, generating an estimate of 10^18 unique clonotypes^37^ In Monoclonal Gammopathies^38^, clonal expansion of B cells expressing a unique and distinct BCR leads to abnormal monoclonal Ig production. In these cases, identifying the sequence of the BCR is useful in diagnosis, risk stratification, and monitoring disease progression.

To improve the detection of BCR clonotypes, the EXaCT-2 assay expanded the Ig variable gene segments (V) to include about 98% of the known repertoire, incorporating diversity (D) and joining (J) intronic regions (Figure 1). Utilizing the bioinformatics tool MiXCR^39^, EXaCT-2 demonstrated effective clonal BCR detection (Figure 3B) on a set of positive control samples assessed with Lymphotrack (Invivoscribe, Inc., USA). For the lymphoma cell line HBL-1^40,41^ MiXCR correctly identified the heavy chain rearrangement at a frequency of 17.6% and various light chain rearrangements (top row). Healthy controls (DLBCL-1, -2, -3 and Urothelial Carcinoma) showed no predominant clonotype and displayed a dispersion of BCR clonotypes at a very low frequency of 2.8% (bottom rows). In two Multiple Myeloma patients, two clonal heavy chain rearrangements at 64.4% (second row), and 60.1% (third row), respectively were detected. The EXaCT-2 assay demonstrated sensitivity with just 1ng of input (Figure S5).

#### Identification of viruses common in hematological and soft-tissue malignancies

Probes for several viruses that are involved in liquid tumors were also included in the assay design, including Epstein-Barr Virus (types 1 & 2 – EBV1/2), Kaposi’s sarcoma-associated herpesvirus (KHSV), Human T-cell lymphotropic virus (HTLV), and Merkel cell polyomavirus (MCPyV). For validation, we collected 14 clinical samples, including one negative control sample and 13 viral positive that included 4 samples with KSHV, and 3 each for EBV, HTLV and MCPyV (Table S10). Apart from the MCPyV specimens, all other samples were initially identified using qualitative PCR. In all viral positive cases, the EXaCT-2 assay was able to detect the presence of each virus (Table S10, see Supplementary Materials for further discussion).

### Depth of Coverage in Cancer Relevant Genes

The EXaCT-2 assay is designed to achieve higher coverage in specific genomic regions highly relevant to cancer profiling. A typical somatic WES assay has an average coverage ranging from 80-120x. However, regions with high-GC content, repetitive sequences, typically may be overly under-represented. Similarly, contamination of normal cells in a tumor specimen reduces the evidence of somatic mutations in the resulting sequencing reads; analogous to sub-clonal events. Thus, we augmented the depth of coverage by selectively adding probes to nearly 1400 cancer genes identified via expert curation (Table S1).

We target to achieve a coverage of about 380-400x for those genes to allow EXaCT-2 to achieve a coverage depth that enables the detection of variants at 5% allele frequency with a probability of 90%^39^. Moreover, an effective 400x coverage is comparable to those provided by targeted panels such as MSK-IMPACT^34^.

Figure 4A illustrates the average coverage in the augmented regions compared to the rest of the assays for 484 samples, including cases and controls. We observed that the expected 400x average coverage in the augmented regions was exceeded (mean = 420, SD=100) and determined that DNA quality has no significant impact on the average coverage (Figure S6). In addition to the augmented regions, we demonstrate the customizability of the assay to allow users to enhance the coverage of notoriously difficult to assay genes, such as CE PA, KRAS, or commonly altered genes such as IDH1 and EGFR (Figures 4B, S7). On average, the EXaCT-2 assay provides 15.8x greater coverage for these genes when compared to EXaCT-1.

**Figure 4:**
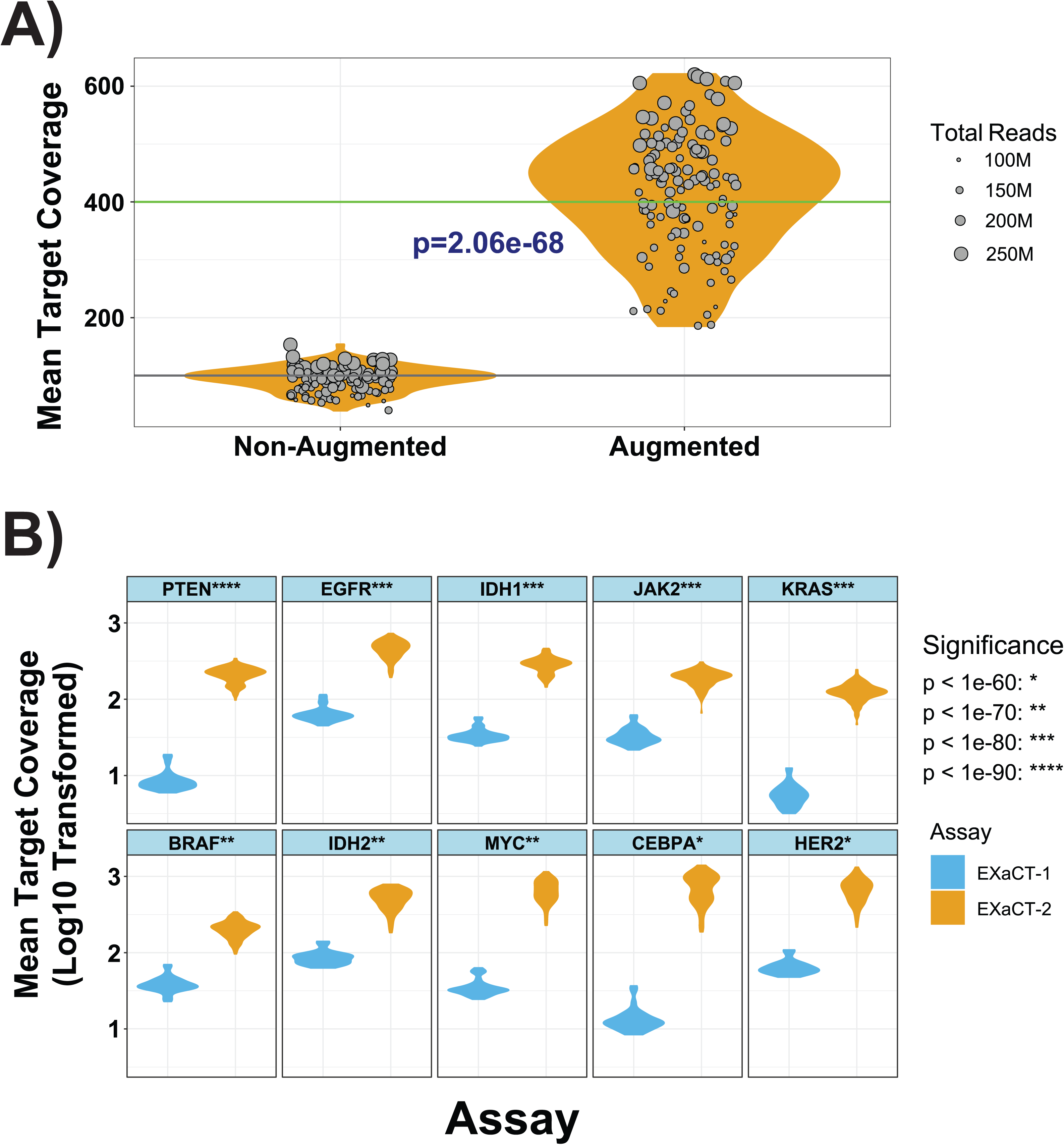
Coverage Analyses. **A) Mean target coverage between augmented and non-augmented regions.** Each dot represents a sample where size is proportional to the Total no. of reads per million. **B) Comparison of mean target coverage on selected cancer genes.** Each plot reports the distribution of log10-transformed mean target coverage on the indicated genes for EXaCT-2 and EXaCT-2.

As an example of this improved coverage, we observed significantly higher coverage for exons 2-5 of the KRAS protein (Figures 5, S8). With the most significant improvement occurring for exon 2, the location of the well-known G12/G13 mutations that are known oncogenic drivers in various carcinomas such as colorectal carcinoma and endometrial carcinoma ^40–43^ (Figures 5A, S8A). KRAS is believed to be one of the most common oncogenic drivers ^44–46^, heavily researched over the past decade^40^. However, the occurrence of this mutation in a highly GC-rich region has made the detection of it with standard NGS panels to be difficult, except when using targeted panels such as the MSK-IMPACT ^34^ assay (Figure 5B). By way of comparison, the augmentation of the EXaCT-2 assay can achieve a coverage nearly equivalent to that of a targeted panel (Figure 5B).

**Figure 5:**
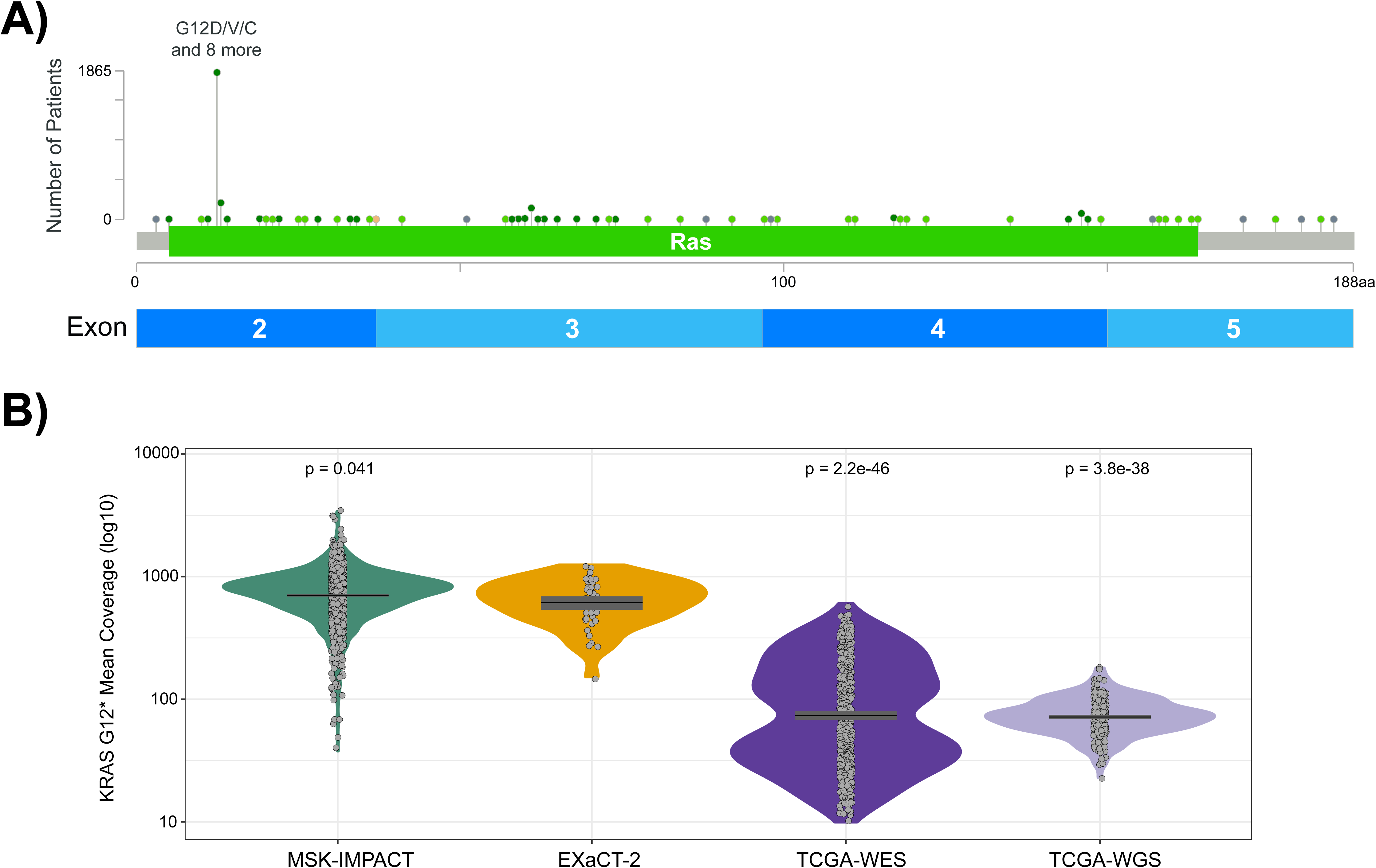
KRAS coverage across protein coding exons. **A) Schematic of the locations of common KRAS mutations from cBioPortal.** The plot shows the frequency of the KRAS^G12*^ mutations. Image generated using a combined cohort of patients with colorectal, pancreatic and non-small-cell lung cancers (see Supplemental Methods). **B) Comparison of the read coverages for the KRAS^G12D^ mutations.** The plot shows KRAS positive samples assayed by EXaCT-2, TCGA Pan-cancer (WES ^47^ and WGS ^48^), and MSK-IMPACT ^34^ (non-EXaCT-2 samples were retrieved from cBioPortal).

Additionally, the EXaCT-2 evaluation of 27 validation samples harboring 9 clinically relevant cancer variants achieved perfect recovery of these mutations with a high degree of variant allele frequencies (VAF) concordance (Figure S9; Pearson correlation: 0.69; p: 5.6e-5).

### Higher coverage in cancer related genes reveals interesting biology

Given this higher coverage in KRAS, we sought to determine if this translated into the EXaCT-2 assay having a higher sensitivity to detect this clinically important mutation. Using the TCGA PanCancer project (whole exome) as a benchmark, we first compared the number of patients that harbored this mutation, relative to the rest of their respective cohorts, limiting the comparison to those samples with cancers that have a known association with the KRAS G12* mutation (colon, lung and pancreatic adenocarcinoma). This evaluation determined that the EXaCT-2 assay had a significantly greater proportion of samples harboring the KRAS G12* mutation than the TCGA (Chi Squared test; p-value = 0.003, table S11). In a subsequent examination, we compared the number of reads carrying the alternative allele (alt reads, henceforth) for mutations with VAF <= 0.2, reasoning that mutations with more alternative reads are more likely to be genuine rather than artifacts. To do this evaluation, we stratified the range of variant allele frequencies (VAFs) from 0-0.15 by increments of 0.05, and determined that in every comparison, the EXaCT-2 assay provides significantly higher alternative read coverage than TCGA WES (Figure 6A).

**Figure 6.**
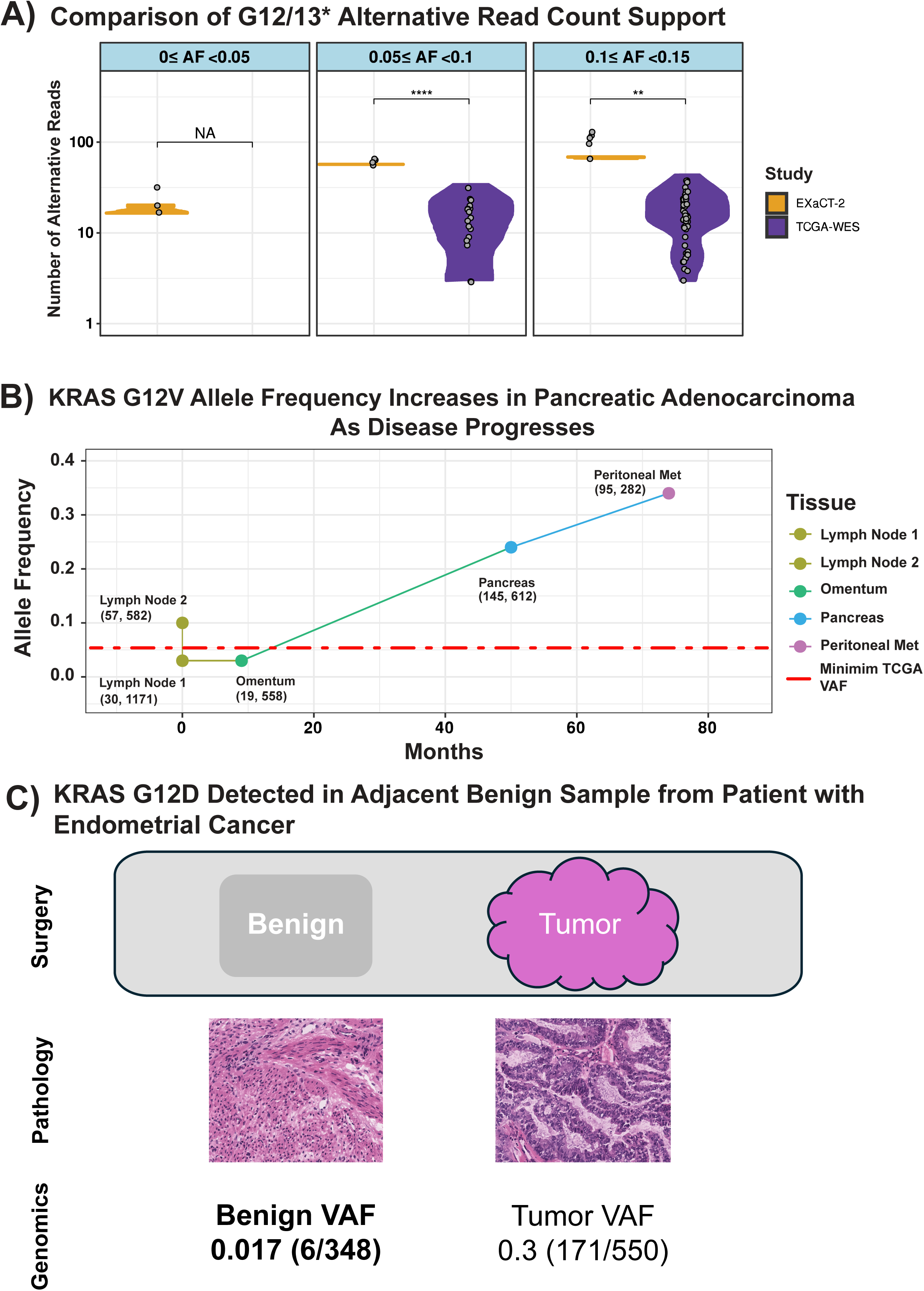
Analysis of KRAS mutations. **A) Number of alt reads per allele frequency.** EXaCT-2 shows higher number of alt reads at the same level of variant allele frequencies compared to TCGA WES data; **B) KRAS G12V mutations in a longitudinal series of 5 PDAC samples.** The early timepoints showed the presence of a sub-clonal population of cell harboring the mutation. **C) KRAS G12D detected in adjacent benign tissue.** EXaCT-2 detects 6 alt reads out of 348 in the benign tissue.

We next sought to determine whether this increased alternative read coverage was generalizable to all the cancer genes whose coverage was augmented in the EXaCT-2 assay. To make the comparison more stringent, we further limited the augmented regions to those that were classified as being problematic by the Genome in a Bottle consortium ^49^, as well as those that possess a GC-enrichment greater than 80% by the UCSC Genome Browser ^50^ (see methods). Limiting our analysis to all mutations that were supported by 15 or more alt reads the EXaCT-2 assay detects a significantly higher number of mutations than the TCGA Pancancer (WES) cohort (Table S12, Chi-square p-value < 2.2e^-16^).

As an initial comparison, we limited our analysis to all mutations that were supported by 15 or more reads carrying the alternative allele (alt reads, henceforth), and determined that the total number of mutations in these problematic regions that were identified when using the EXaCT-2 assay was significantly greater than those that were identified within these regions in the entire TCGA Pancancer (WES) cohort (Table S12, Chi-square p-value < 2.2e^-16^). One can visualize this difference when one compares the distribution of the mutational VAFs that are provided by the EXaCT-2 assay with those generated from standard sequencing platforms like that which was used by the TCGA PanCancer project ^47^ (Figure S9). In this comparison, the critical difference occurs in the lower tails of these distributions which demonstrates that EXaCT-2 assay has the sensitivity to capture more sub-clonal mutations than is possible with standard sequencing assays, and with far greater confidence.

For example, we identified 10 samples in our cohort that possessed a KRAS G12* mutation with allele frequencies less than or equal to 0.1. Of these, 5 samples belong to a 69-year-old, Caucasian female patient with pancreatic ductal adenocarcinoma (PDAC) which were collected over the span of more than 6 years^54^. EXaCT-2 detected the G12V mutation, that is well-known to be associated with this disease, in all of them ^51–53^. Importantly, when this case was examined previously, using a pyrosequencing assay specifically designed for KRAS ^55^, only the 2 later samples were determined to possess the KRAS G12V mutation ^54^. Indeed, EXaCT-2 assay detected this mutation with the allele frequency progressively increasing from 0.03 to 0.34 over this time period and treatment protocol (Figure 6B).

Similarly, in another example, the KRAS G12A mutation was observed in 2 samples that were collected from a patient with endometrial cancer, one of which was from the tumor, itself, while the other was from a proximal normal tissue (Figure 6C). In this instance, the mutation was observed with an allele frequency of 0.31 in the tumor sample, while it was also observed in the proximal normal with an allele frequency of 0.017, indicating the presence of cells harboring this mutation. This could be explained by pockets of tumor cells in the benign tissue or by aging-related, somatic mosaicism within this tissue^56–59^.

Finally, the EXaCT-2 evaluation of 27 validation samples harboring 9 clinically relevant cancer variants achieved perfect recovery of these mutations with a high degree of variant allele frequencies (VAF) concordance (Figure S13; Pearson correlation: 0.69; p: 5.6e-5).

## Discussion

Next-generation sequencing (NGS) has revolutionized cancer research and treatment by identifying a broad spectrum of genomic alterations that drive the disease. This has facilitated the development of targeted treatments, thus improving survival and reducing toxicity for various tumor types. Targeted assays excel in precision, detecting sub-clonal mutations in a limited number of known cancer driver genes. However, broader assays have also revealed a long-tail of cancer mutations, with only a few genes exhibiting common alterations ^60^. Given this complexity of the cancer landscape, an accurate and comprehensive characterization of clinical samples is required. WES assays detect coding mutations on all genes but cannot accurately identify intergenic events. WGS assays cover the broadest genomic territory but have limited coverage especially in GC-rich regions and remain expensive, especially considering the hidden costs of processing and long-term data storage.

EXaCT-2 enhances *depth* of coverage based on known cancer genes and expand the *breadth* by interrogating intronic and intergenic regions to detect common rearrangements and improve copy number boundary detection. Additionally, it examines the presence of viruses, expands the evaluation of the BCR repertoire, and is customizable, allowing rapid adaptation to new discoveries, as demonstrated by the increasing coverage of GTF2I in thymoma or CEBPA (Figures S5, S10)^61^.

Global metrics such as tumor mutation burden (TMB) and microsatellite instability (MSI) have emerged as crucial prognostic biomarkers for patient outcomes and responses to immunotherapy^19–22, 65–68^. Whole-exome sequencing (WES) is considered the gold standard for accurately estimating TMB.^69, 70, 18, 71,72^ EXaCT-2, which evaluates both coding and non-coding regions, can further explores correlations between TMB and MSI (Figures S11, S12), confirming that higher TMB does not necessarily correlate with higher MSI.^73, 74^

The higher depth of EXaCT-2 has enabled the capacity to detect critical mutations, such in the case of the longitudinal samples at low levels (Figure 6B), providing clinicians more informed treatment choices at earlier stages of the disease than is possible with standard whole exome assays.

Having established the biological insights yielded by EXaCT-2, we also evaluated the effort and costs involved in running the assay. EXaCT-2 requires 3 days for library preparation and a sequencing depth of 125M reads per sample, comparable with targeted panels and only ~15% more expensive than TSO500 and ~24% cheaper than EXaCT-1 (see Table S14 for a full comparison). The bioinformatics pipeline for EXaCT-2 is computationally intensive, but with optimized computing and parallel processing, the total turnaround time for a 12-sample run is approximately 10-12 days. Limitations are the need for matched tumor/normal specimens, increasing costs and logistical complexity, and the ability to detect only selected rearrangements (n=66 genes). However, the former is common to other targeted assays and the latter is comparable to the RNA component of the TSO500 assay (n=55 genes) ^34, 75^. As a DNA-based assay, it cannot determine if rearrangements are expressed, so complementing it with a transcriptome-based assay provides a comprehensive molecular picture.^76^

Overall, EXaCT-2 combines the features of a clinical-grade assay with the capacity for novel discovery, by balancing the strengths of targeted panels and WGS assays and offering a more comprehensive view of tumor alterations—particularly important given the broader genomic diversity observed in non-European populations, which remains underrepresented in current targeted panels^13, 62–64^.

## Methods

### Sample Collection, DNA Preparation, Library Construction and Sequencing

Samples from 120 cancer patients were collected (IRB1305013903, IRB0107004999, IRB1008011221, IRB1011011386). H&E-stained slides were annotated by board-certified pathologists and tumor DNA was excised from fresh frozen tissues or FFPE. Germline DNA was obtained from blood, buccal swabs, or benign tissue. DNA was isolated using Promega Maxwell 16 (Promega) and evaluated using Qubit dsDNA HS Assay (ThermoFisher Scientific) and TapeStation System (Agilent). DNA from 2 Multiple Myeloma patients was derived from CD8+ bone marrow biopsies (IRB10004608). FFPE from 28 patients for fusions and viral content, and DNA from 27 patients for clinical mutations studies, were provided by the WCM Clinical Lab; HBL-1 DNA by Dr. Tam, WCM HemOnc. DNA with Integrity Number (DIN) <4 was repaired using NEBNext FFPE DNA Repair MIX M6630 (NewEngland BioLabs). Libraries were prepared with 50ng (DIN>-4) or 200ng (DIN<4) of DNA using SureSelect Enzymatic Fragmentation and XT Low Input kits following manufacturer instructions (Agilent, Cat 5191 and G9703A). Indexed libraries (~1500ng) were hybridized with 5µl of the EXaCT-2 Target Enrichment probes (Agilent, Cat G9496D). EXaCT-2 probes were designed using high-density tiling (5X), with a booster probe set for cancer regions with <50X coverage and an augmented probe set for a subset of these gene. Probes are approximately 120bp size, which also is about the median exon size of human genes.^77^ EXaCT-2 composition: SureSelect V6 + COSMIC (1X), Non-Augmented Regions (1X), Augmented Targets (5X), Booster Probe Set (5X), CEBPA (300X), and Viral Genomes (1/30X). Libraries obtained were sequenced for 150 cycles on a pair-end flowcell (Illumina NovaSeq6000) to obtain ~125M read per sample.

### Bioinformatics processing

Data were processed using Illumina’s RTA and bcl2fastq software to generate and demultiplex FASTQ files. Quality Control analysis of the sequencing run was performed with fastp^78^. Reads passing Illumina’s purity filter are adapter trimmed and aligned to a modified version of the GRCh37 human genome that includes the viruses included in the assay. Quality control analysis is performed upon the result BAM files, and the matched tumor-normal samples are analyzed with SPIA^79^ to ensure that the samples are from the same individual.

The tools used for the different analyses for each matched tumor-normal sample pair are listed hereafter (details provided in Supplementary Methods): An updated version of the MC3 algorithm (SNV & Indel Identification)^80^, and Nirvana (annotation)^81^; CNVkit (SCNA Identification)^82^; MSISensor-Pro (Microsatellite instability)^83, 84^; Delly (Rearrangement Detection)^85^; in-house calculation for Tumor Mutation Burden (TMB)^72, 86^; MiXCR (Immuno-globulin (Ig) rearrangements)^87^; in-house tool for Viral Detection.

## Data Availability

All data produced in the present study are available upon reasonable request to the authors

## Acknowledgements

We acknowledge help from the following institutions and individuals. Giuseppe Giaccone, MD/PhD, for the GTF2I and MTAP customization discussion. The Genomics and Epigenomics Core Research Facility at Weill Cornell Medicine for the library preparation and sequencing. The Englander Institute of Precision Medicine for funding this project.

