## Supplementary Materials and Methods for "EXaCT-2: An augmented and customizable oncology-focused whole exome sequencing platform"

### Supplementary Results and Methods

##### Enhanced precision in detecting boundaries of somatic copy number alterations (SCNAs)

To explore the advantages that the expanded set of probes provide to the identification of SCNAs, we extended the comparison performed above to 4 clinical samples that were analyzed by both EXaCT-1 and EXaCT-2. As a first pass, to ensure that both assays produced similar results for these matched samples, we first performed a gene-wise comparison of the results from the analysis from each assay (Table S3) and determined that the results were statistically similar (Chi-Square p < 0.00001).

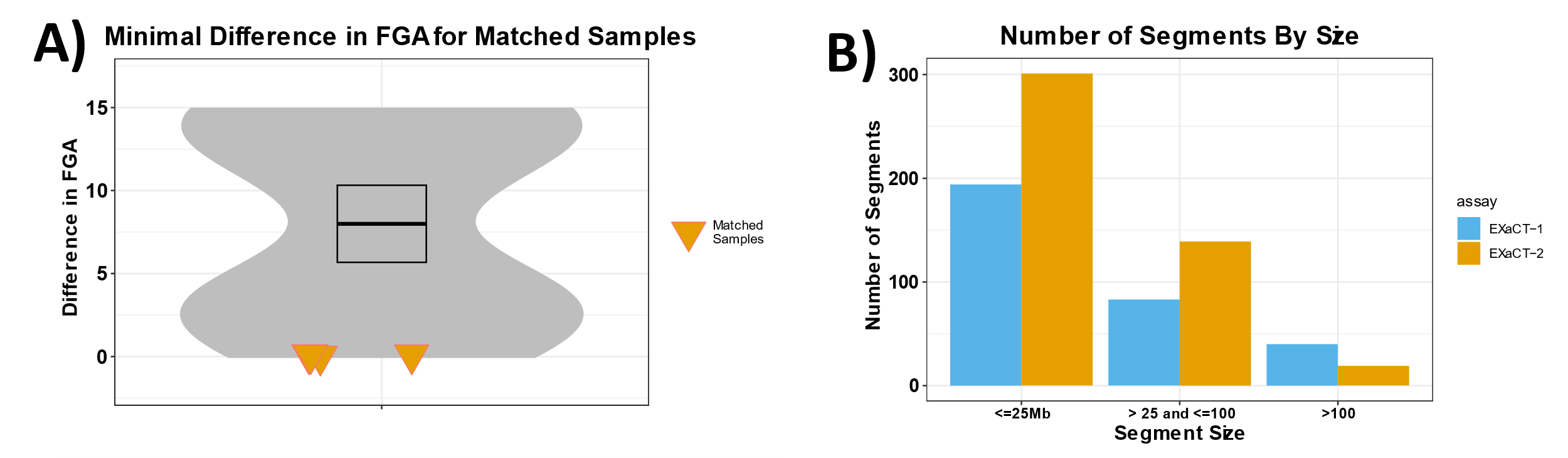

**Figure S1: Comparison and analysis of SCNA segments across four matching WES assays**. **A)** Difference in the total Fraction of Genome Altered (FGA): No major difference is observed in FGA between matching EXaCT-1 and EXaCT-2 samples; matched samples are highlighted and compared against a simulated null that was generated by calculating the difference for all possible pairwise comparisons of the samples. **B)** Grouping segments based on their size: As number of segments suggest, EXaCT-2 shows a higher prevalence of smaller segments (<25Mb) and lower number of larger segments (>100 Mb) compared to EXaCT-1.

In addition to the overall gene-wise similarity of these results, we observed that the total fraction of the genome altered (FGA) by an SCNA was nearly unchanged between the two assays (p=0.59; Figure S1A, Table S4). In contrast, however, the total number of inferred segments was considerably higher in the results from the EXaCT-2 assay, indicating that the EXaCT-2 assay might provide for finer granularity of the segmentation generated by the SCNA analysis (p=0.056; Figure 2C-Top, Table S5). To further examine this hypothesis, the segments generated from the two assays were stratified by segment length and compared. While there was no statistical difference between the mid-sized segments (those between 25-100Mb), the EXaCT-2 assay produced significantly fewer large (>100MB) and small segments (<25Mb), indicating that the EXaCT-1 segments were both over- and under-segmented, in comparison to those from the EXaCT-2 assay (Figure S1B).

Given these results, we performed a systematic analysis and comparison of the segmentations generated by the two assays (Figure S2). As a first pass, we were interested in the number of segments that were identified by both assays, as well as those which were unique to each assay. When using an extremely simplified definition of concordance as being any region in one assay where at least half of it overlapped with one or more regions from the other assay, we discovered that on average 98.1% of all segments from EXaCT-1 overlapped with those identified in EXaCT-2, and 95.8% of all EXaCT-2 segments overlapped with those identified with EXaCT-1 (see Methods for details on how SCNAs were identified for each assay). Note, however, that even when using a more stringent threshold by requiring 95% overlap between the segments, these proportions only drop to 87.8% and 89.0%, respectively. Note, that with this simplified definition of concordance, we do not consider inferred copy number for reasons that we will discuss further below.

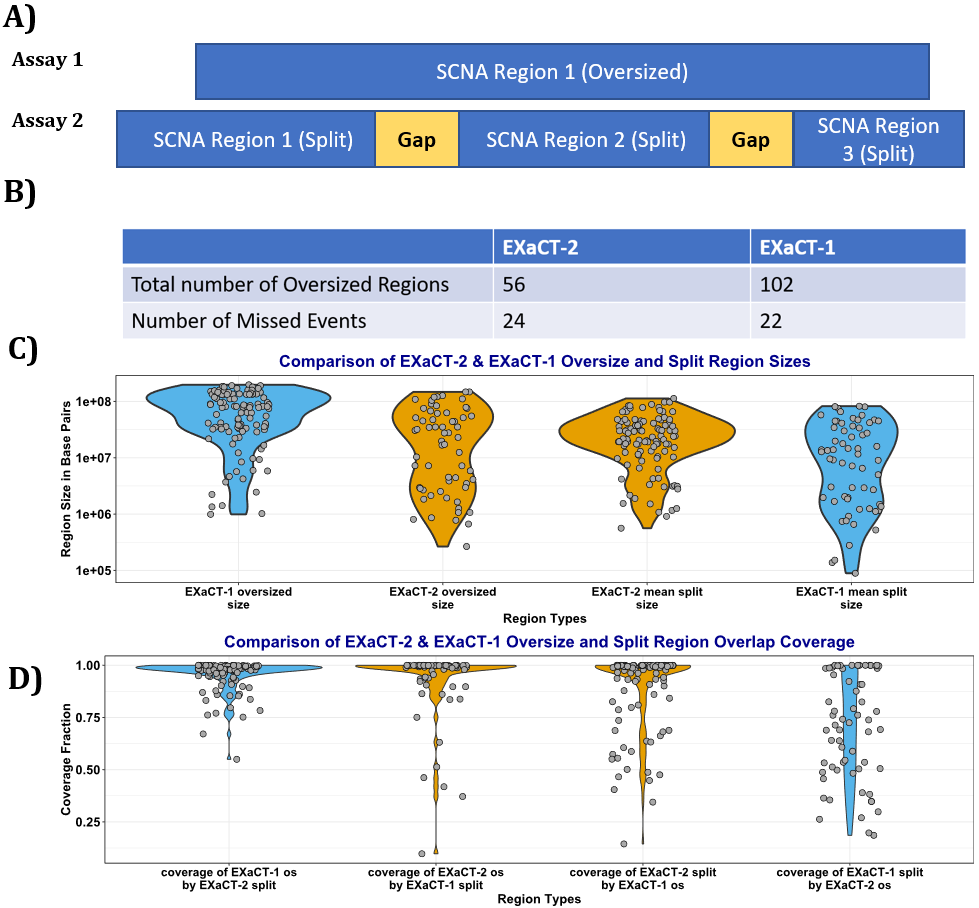

**Figure S2:** A) Schematic of the oversized regions from one assay, and the split regions from the results of the other, where in a given region Assays 1 and 2 identify somatic copy number alterations and segments, but Assay 1 has generated a region that was decomposed into smaller regions by Assay 2. **B)** A comparison of the total number of oversized regions for EXaCT-1 and EXaCT-2, along with the total number of discordant events (SCNA events that were identified in the results from one assay, but not identified in the results of the other assay). **C)** Comparison of the sizes of the **oversized** and **split** regions from the EXaCT-1 and EXaCT-2 regions. In this panel, we demonstrate that the EXaCT-2 oversized and split regions are similar in total size, while the EXaCT-1 oversized and split regions are statistically larger and smaller than each other, as well as their respective classes from the EXaCT-2 assay, indicating that the segmentation from the EXaCT-1 assay is both under- and over-segmented, respectively, relative to the segmentation from the EXaCT-2 assay. **D)** Comparison of the reciprocal overlapping coverage of the **oversized** and **split** regions from the 2 assays. In this figure, we demonstrate that the oversized regions from EXaCT-1 are nearly perfectly covered by their respective split regions from the EXaCT-2 assay. Similarly, we demonstrate that the EXaCT-2 oversized regions are nearly perfectly covered by their corresponding EXaCT-1 split regions, however, the coverse is not true. Instead, the EXaCT-1 split regions are significantly less well-aligned and less concordant than the results from EXaCT-2.

Of these overlapping regions, we next sought to identify the number segments that were mutually and uniquely identified by both assays, meaning segments from one assay that mutually and uniquely overlapped segments from the other assay. We further limited these matches to those where the overlap between the 2 segments was at least 80% for at least one segment, and they had concordant copy number rations (both concordantly showed amplification or deletion, even if insignificantly). Given this definition of concordance, the overall average number of matching segments was 23.5. However, with a standard deviation of 28.8, these results indicated a high variability in the number of matching segments.

Upon investigation, it was determined that this high variability resulted from 2 samples that had almost no shared segments (n <=1 for both). For both samples, the EXaCT-2 segmentation had nearly twice the number of segments from the EXaCT-1 segmentation, or higher, indicating that the EXaCT-1 assay led to an under-segmentation of these samples. Admittedly, some caution should be taken, given the small samples sizes, but even when these 2 samples were excluded, the average number of shared segments was 46.5, with a standard deviation of 19.1, which still represents a small number of segments for either assay for the remaining 2 samples.

Given these indications of under-segmentation from the analyses with EXaCT-1, we next sought to explore and quantify the degree of under- or over-segmentation from either assay. To do this, we first introduce a couple of terms. We define an **undersegmented segment** as any segment from one assay that overlaps with more than one segment from the other. Conversely, we define a **split segment** as a segment from one assay that, along with other adjacent split segments, overlaps with an undersegmented segment from the other assay (Figure S2A).

Overall, we observed that the number of undersegmented EXaCT-1 regions was significantly greater than the number of undersegmented EXaCT-2 regions (p=0.042). Likewise, the number of EXaCT-2 split regions was also significantly greater than the number of split regions (p=2.25e-8). This was also reflected in the average size of these regions (Figure S2B), where the EXaCT-1 undersegmented regions were significantly larger than the EXaCT-2 undersegmented regions (p=4.51e-7), while there was no statistical difference in the size of the EXaCT-2 undersegmented and split regions (p=0.15). Ironically, we also noticed that the EXaCT-1 split regions were significantly smaller than any of the others (p=7.26e-4 when compared with all of the EXaCT-2 regions). We also were struck by the reciprocal overlap of the undersegmented and split regions, e.g. how much of an undersegmented EXaCT-1 region overlaps with split regions from the EXaCT-2 assay, and vice versa (Figure S2C). With the exception of the EXaCT-1 split regions, the median overlap was 0.99.

##### Fusions

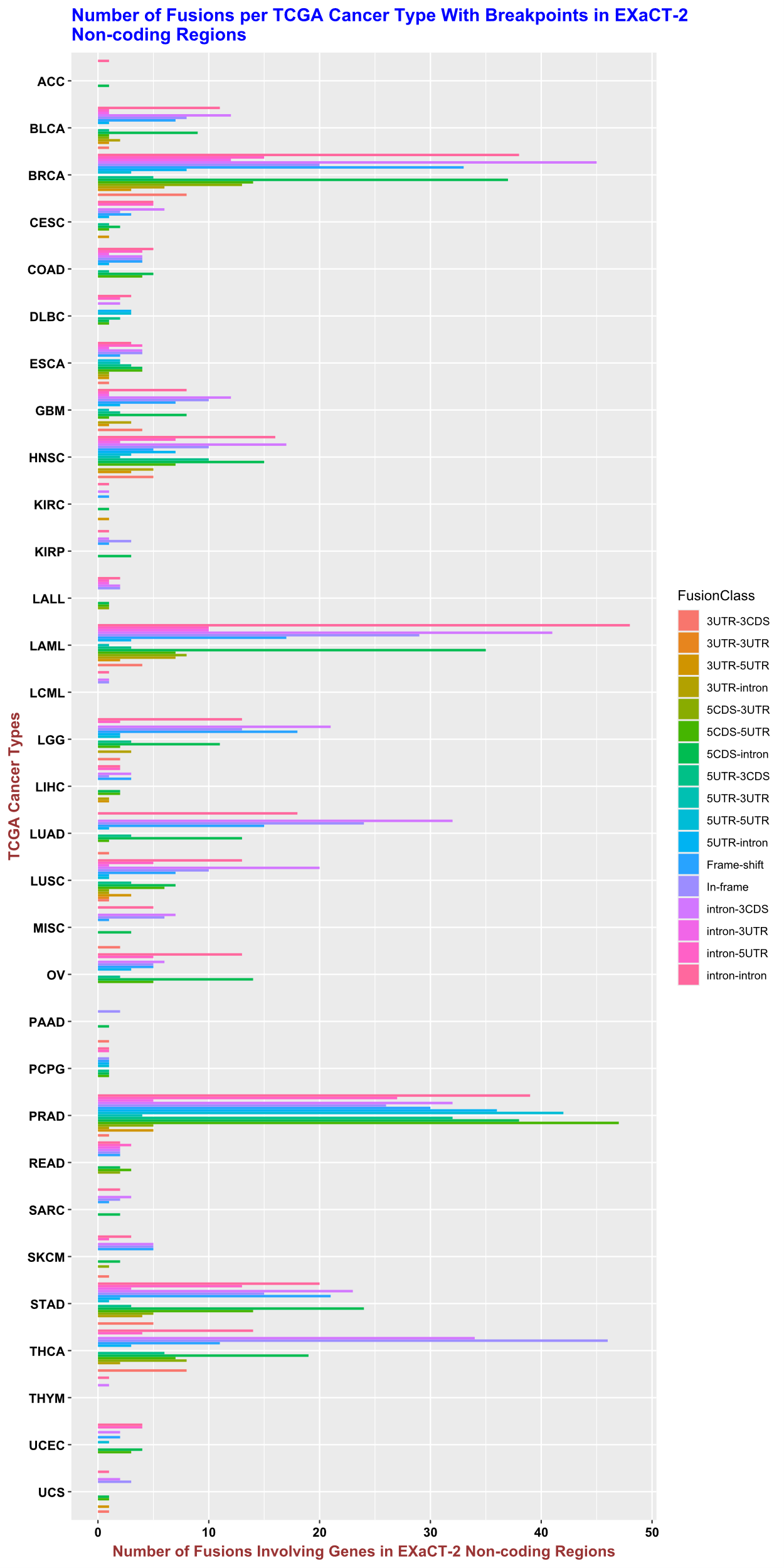

**Figure S3: Broad Spectrum of Fusions Targeted by EXaCT−2.** We display the broad spectrum of known fusions that have at least one breakpoint in the non-coding regions that are targeted by the EXaCT-2 assay. As shown, fusions are separated by cancer type (using TCGA acronyms) and fusion class, as defined by the source that provided the list of fusions. This list of fusions was downloaded from FusionGDB v.2.0 ^1^, and limited to those which have at least one (1) breakpoint that occurs in the non-coding regions that are targeted by the EXaCT-2 assay. To prevent double-counting fusions, we define those that share the same breakpoints (same chromosomal coordinates for both breakpoint1 and breakpoint2) as being duplicate fusions, and limit the set to only including one representative of that fusion.

###### NTRK2-KANK2 Rearrangement

As a negative control, we were also provided 2 samples that were identified to possess fusions involving Neurotrophic Receptor Tyrosine Kinase 2 (NTRK2—KANK2 and NTRK2—AFAP1), as none of the genes involved in these fusions were included in the set of 66 genes that were targeted for fusion detection, as described above. While EXaCT-2 was unable to recover both of these fusions, it was able to recover the NTRK2-KANK2 fusion as the breakpoint for the KANK2 gene occurred in the exon padding region on the 3’ side of the fourth exon (Table S7 and Figure S2).

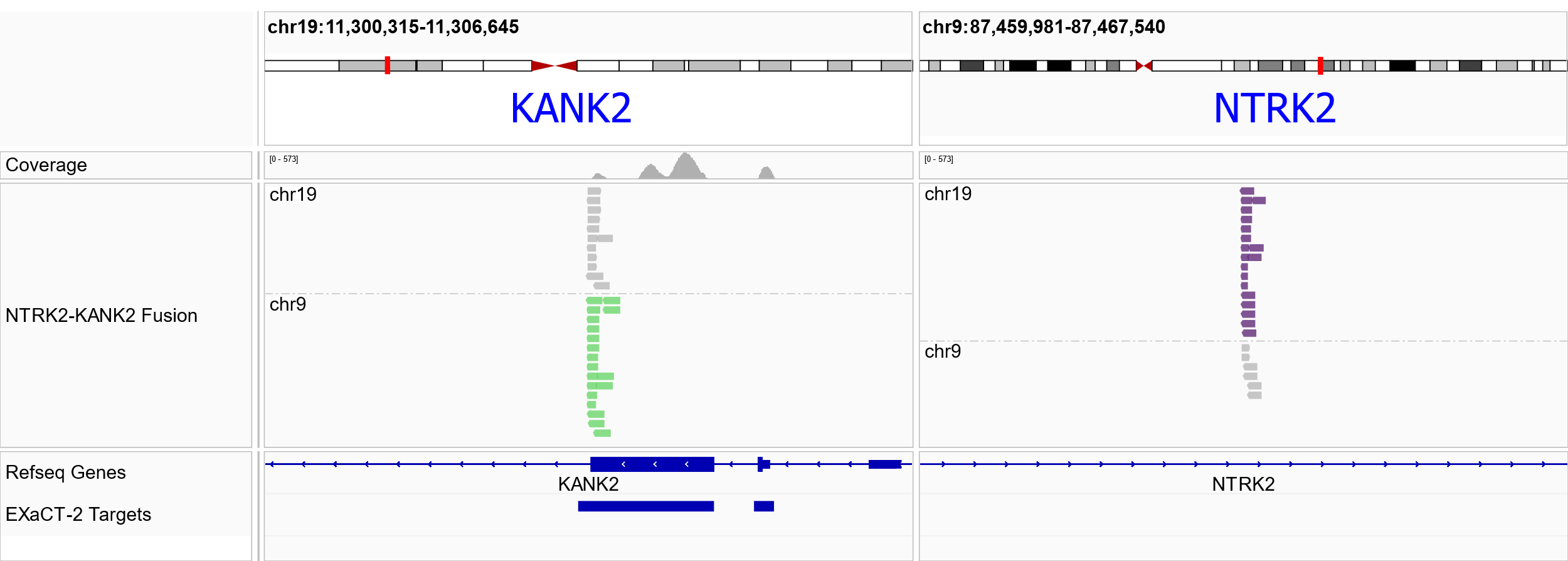

**Figure S4: IGV representation of the NTRK2-KANK2 fusion that was discovered by EXaCT-2.** Similar to figure 5B, we show the reads supporting a putative fusion involving NTRK2 and KANK2. In the left-hand side of the figure, shown in green are the reads whose mate pairs align to chromosome 9, while, in the right-hand panel, reads whose mate pairs align to chromosome 9 are displayed in purple. Note, that the reads in the NTRK2 region don’t align to an EXaCT-2 target region, nor an NTRK2 exon (regions shown along the bottom of the figure); while, in contrast, the reads in the KANK2 region align to a KANK2 exon boundary that is captured by the EXaCT-2 assay.

##### Sensitivity of EXaCT-2 in detecting immunoglobulin rearrangement

To determine the EXaCT-2 sensitivity to detect clonal rearrangements we used the HBL-1 cell line at decreasing dilution levels (10ng, 5ng, and 1ng). We focused our analysis upon all three rearrangements observed in the native HBL-1 cell line. As illustrated in Figure 7, the presence of IGLV2-5 and IGLJ3 clonotypes with 10ng the IGLV2-5/IGLJ3 clonotype was observed at 55.9%, but dropped to 23.3% in the 5ng sample, and 3.3% in the 1ng sample, confirming the capacity of the EXaCT-2 assay to detect clonotypes in of both high and low concentrations. Furthermore, clonotype rearrangement, IGKV3-20 and IGKJ3, was also observed at decreasing read frequencies; HBL-1 10ng (22.9%), HBL-1 5ng (10.8%) and HBL-1 1ng (2.9%). Lastly, the heavy chain rearrangement, IGHV4-34, IGHD4-17 and IGHJ4 was detected only at 10 ng (17.6%) and HBL-1 5ng (5.9%). This rearrangement was not predominant at 1ng, yet it was detected among other rearrangements in the patient. This indicates that the assay is sensitive, and once a clonal rearrangement is detected, it can be followed up as a biomarker.

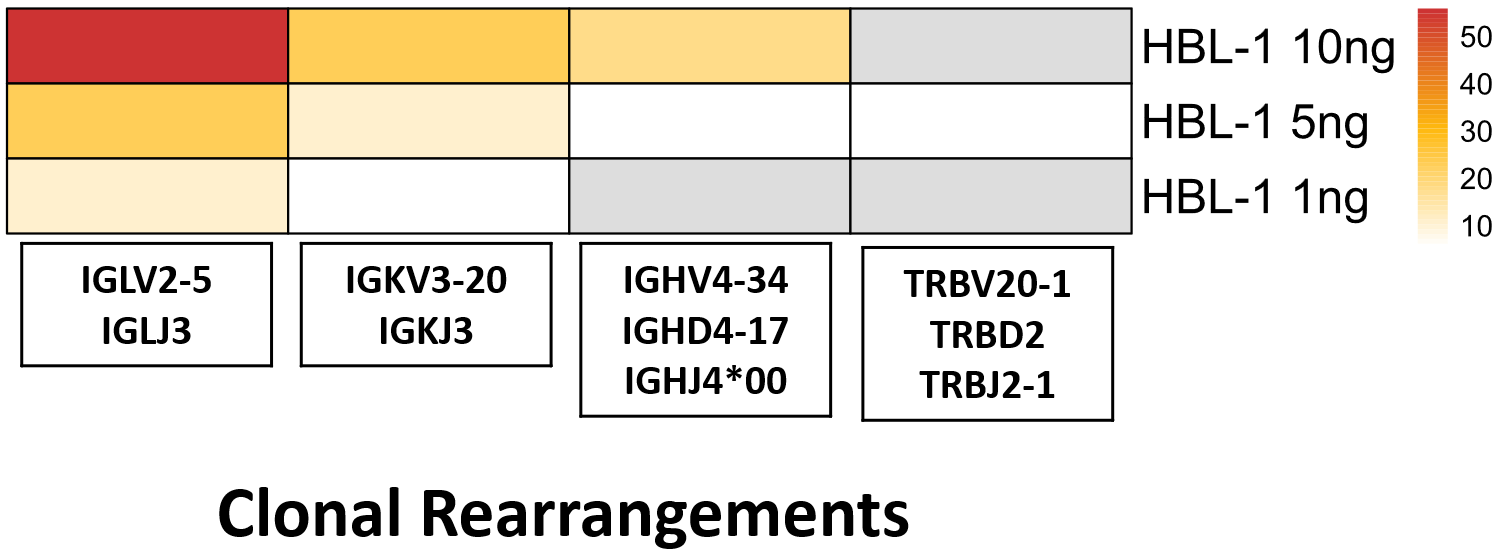

**Figure S5. Limit of detection of immunoglobulin rearrangement.** All the highlighted rearrangements in the box have a reads frequency of >= 5%. The color scheme follows the pattern depicted in Figure 6, where red indicates the highest frequency, followed by orange and then yellow. White represents the lowest frequency, while gray signifies a read frequency of 0.

##### Discussion of detection of carcinogenic viruses by EXaCT-2

As described in the main text, probes for several viruses that are involved in liquid tumors (EBV, HTLV, KSH and MCPyV ) were included in the assay design, and their capacity to detect these was tested with 14 clinical samples (Table S10). With the exception of the MCPyV specimens, all other samples were initially identified using qualitative PCR.

In contrast, the MCPyV samples were from specimens with a diagnosis of Merkel cell carcinoma (MCC) that hadn’t been previously tested by other molecular assays prior to our analysis with the EXaCT-2 assay. It is well-established that MCC, a rare, aggressive skin cancer that is driven by either ultraviolet-induced DNA damage or MCPyV infection ^2,3^. As such, EXaCT-2’s capacity to detect MCPyV can and will help inform the treatment options chosen by clinicians for their patients, which is critical as it has been shown that MCPyV-negative patients are 40% more likely to experience a recurrence ^4^.

As described in the main text, the EXaCT-2 assay was able to detect the presence of each virus in all positive cases, and in with some of these, was able to detect dual infections, with EBV being the most commonly co-infected virus, which we attribute to the commonality of EBV infections population-wide (Table S10). In addition to the EBV co-infections, both KSHV and MYCPyV were also detected in trace amounts in samples not previously known to be infected with them (n = 5 and 6 coinfections, respectively, including the one negative control sample). However, in each case of these incidental detections (for lack of a better term), fewer than 40 reads were mapped to these off-target viruses, in contrast to the positive control cases, where the number of reads usually exceeded 1000. Admittedly, this is a heuristic threshold, however at the time of the writing, it is unclear from the literature what are appropriate thresholds to use for NGS testing, especially as viral length will impact the number of reads that are detected. As we expand the use of the EXaCT-2 assay, we may be able to identify a statistically rigorous threshold, however, we note that it has been shown that some viruses, such as EBV, may be present in the host genome, even in low frequencies. ^5–7^. For this reason, we report all results to clinicians to allow them to determine the appropriate course of action.

##### Depth of Coverage

The depth of coverage provided by EXaCT-2 is robust to DNA quality, as measured by DNA Integrity Number (DIN).

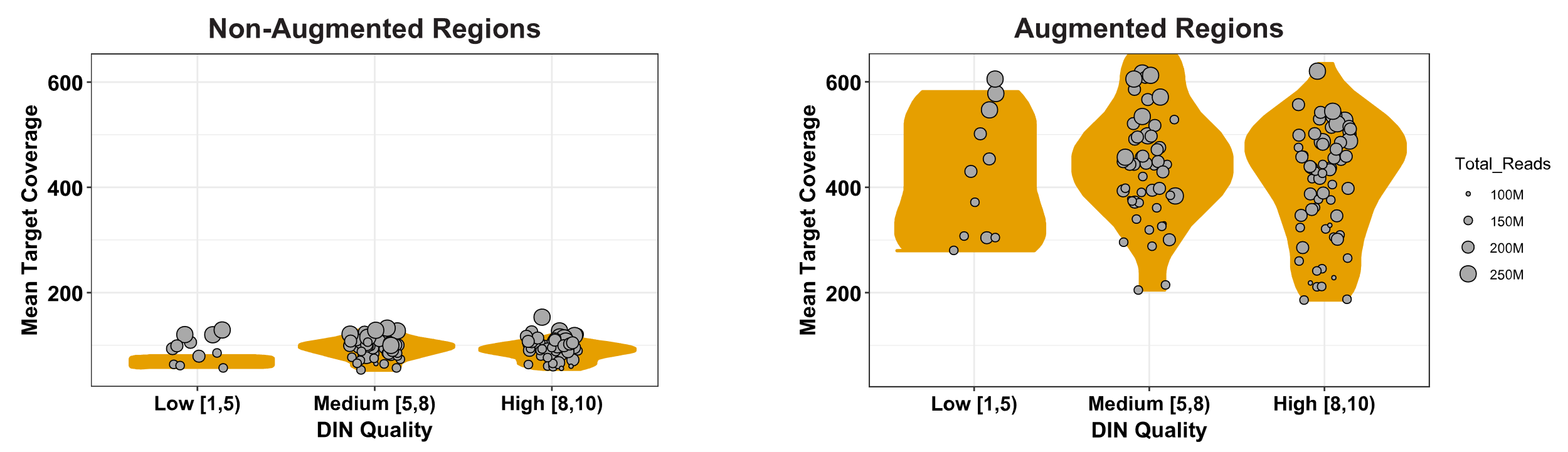

**Figure S6: Comparison of mean target coverage and DNA quality.** Samples are classified by DNA Integrity Number in 3 categories: Low, Medium, and High. Left: non augmented regions; right: augmented regions. Surprisingly, DIN quality has no significant effect on the average coverage provided by the EXaCT-2 assay for either the augmented or standard (non-augmented) regions.

##### CEBPA coverage: improved coverage for GC-rich regions

Figure S7 demonstrates the far higher coverage of a region in the CEBPA gene with greater than 80% GC content. Whereas the matching EXaCT-1 sequencing for this sample only had a maximum coverage of 87 reads in the coding region, the corresponding region for this sample in EXaCT-2 had a maximum coverage of 3506 reads from the EXaCT-2. Furthermore, there are multiple segments within the CEBPA coding region in which the EXaCT-1 assay had no or minimal (<10 reads) coverage.

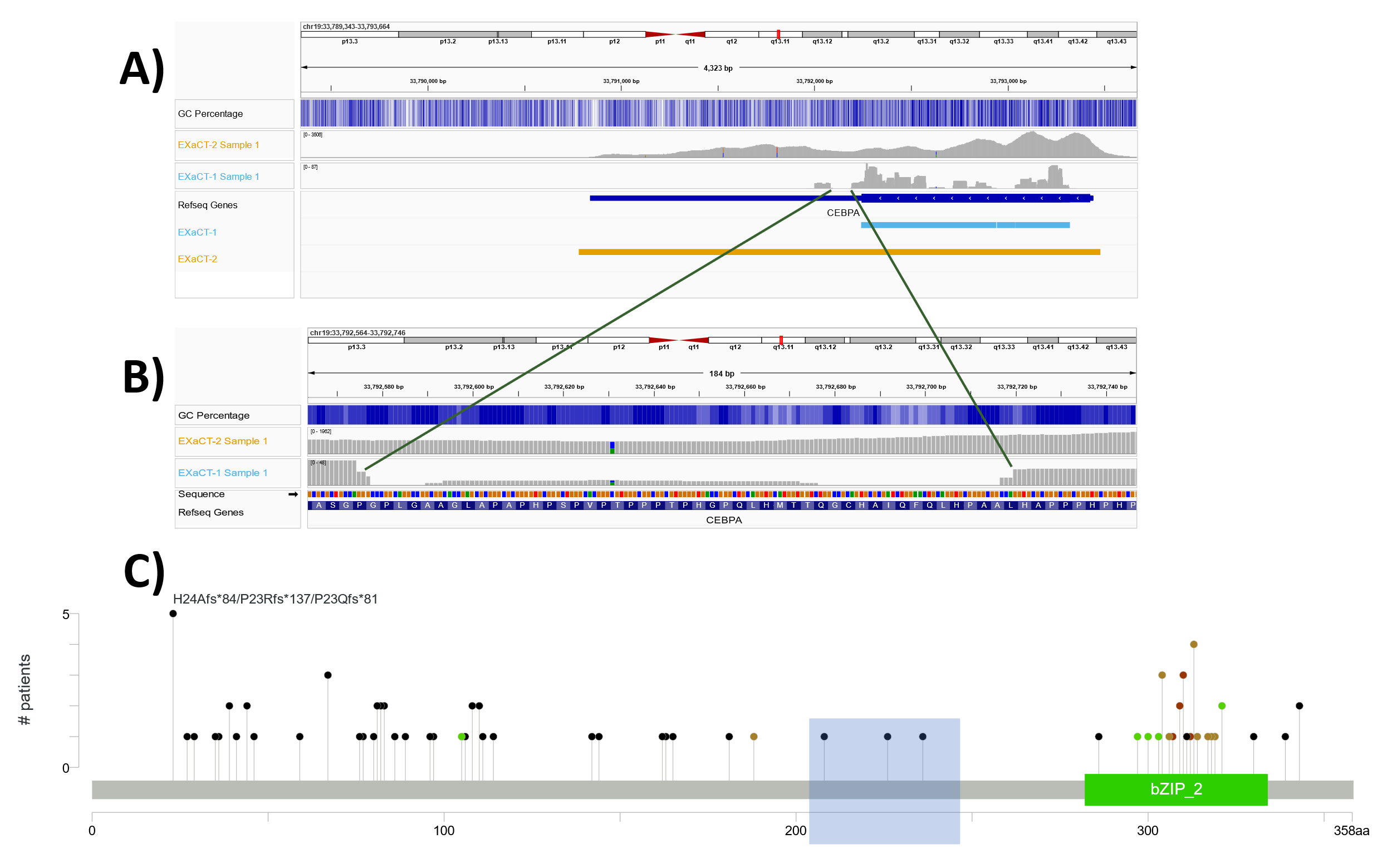

**Figure S7: Enhanced coverage in high GC content regions:** A) EXaCT-2 sample has a read count of ~5400 with GC content (~80%) in CEBPA gene as compared to EXaCT-1 having a read count of ~140. B) We highlight a small segment of the CEBPA exon with extremely sparse coverage in EXaCT-1, and compare the coverage to that which was achieved with EXaCT-2. C) Lollipop plot of known mutations for CEBPA using a cohort of patients with Acute Myeloid Leukemia, collected from public sources. The GC-rich region with low coverage that is discussed in the text is highlighted in blue.

Focusing on one such region, a GC-rich 143 nucleotide long segment that represents amino acids 201-248 in CEBPA (chr19: 33,792,577- 33,792,719), we observed that the average coverage provided by EXaCT-2 in this region for this sample was 1073 reads, whereas the maximum coverage provided by EXaCT-1 was only 17 reads, with an average of 5.5 reads and 52 positions with no coverage whatsoever (Figure S7B). Interestingly, when examining public databases of known cancer mutations in patients with Acute Myeloid Leukemia, there appears to be a sparsity of known mutations in this poorly-covered GC-rich region (n=3), suggesting that this may be a general problem for the field, regardless of the assay (Figure S7C). When the read coverage of these mutations is examined, the number of reads carrying the alternative allele is, on average, only 5.3 reads with a mean total coverage of 20 reads, indicating that even in the case of these mutations, the read support was relatively limited.

##### KRAS

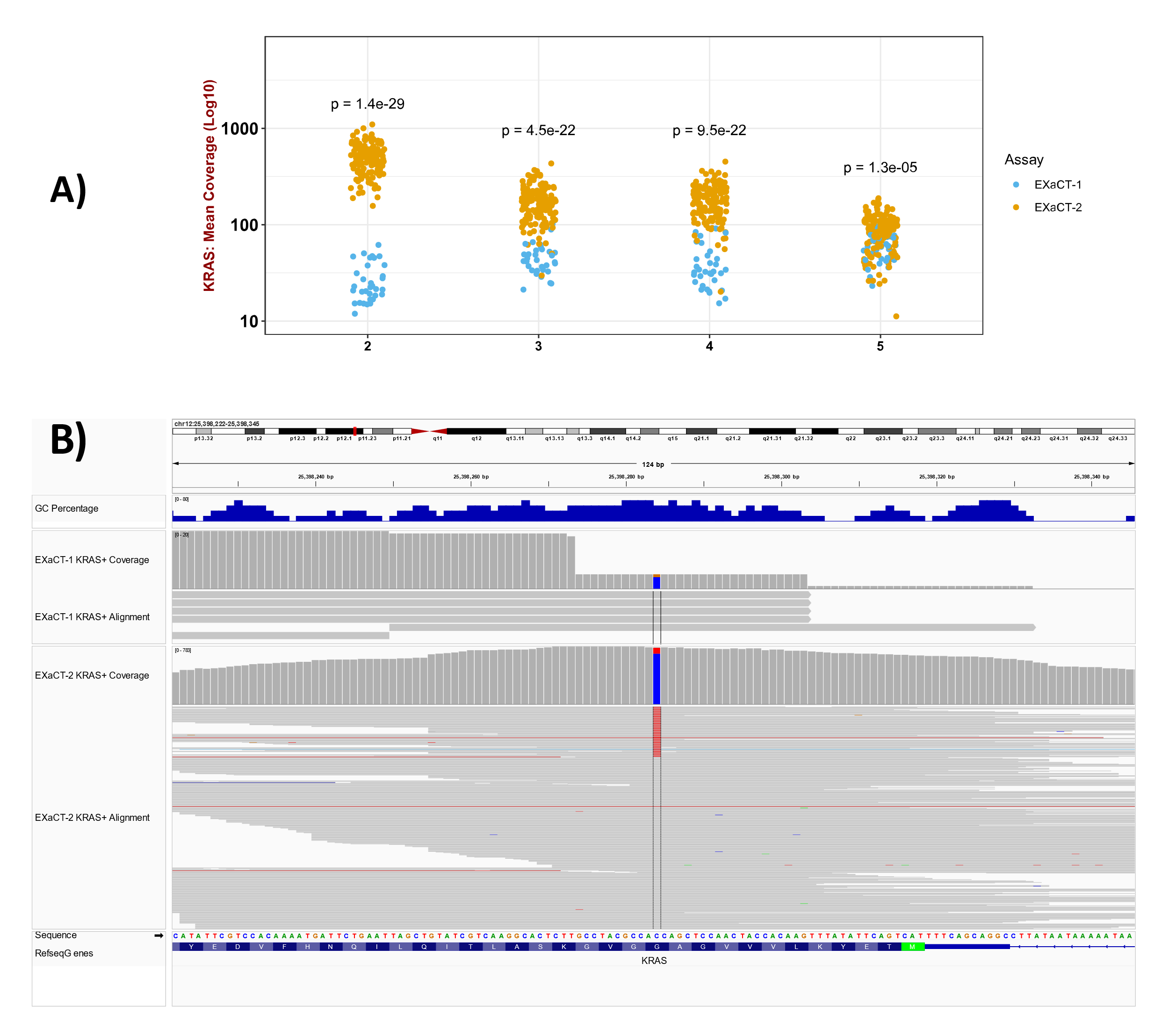

Figure S8**: KRAS coverage across protein coding exons.** **A)** The average read coverage, per exon, of the KRAS gene is shown for samples analyzed by the EXaCT-2 assay, as well as that produced by standard WES assays, such as the EXaCT-1 assay. As shown here, exon 2, location of the well-known G12D mutation, has significantly higher coverage in EXaCT-2 compared to EXaCT-1. **B)** IGV Visualization of KRAS G12D positive samples assayed by EXaCT-1 and EXaCT-2 samples. In the top panel, we display the coverage and alignment tracks for an EXaCT-1 sample that was positive for the well-known G12D mutation, in which the coverage was only 4 total reads (with 1 carrying the alternative allele). By way of comparison, in the bottom panel, we display the coverage tracks for an unrelated G12D+ sample that was assayed by EXaCT-2. While the allele frequency in this sample was only 0.11, the number of reads with the mutant allele was 86 (out of 772 total reads covering this position).

Figure S8A demonstrates the significantly higher coverage that we observed between the EXaCT-2 and EXaCT-1 assays for the coding exons of KRAS. To further explore this, we compared the coverage and read alignment of a KRAS^G12D+^ sample that had been assayed by EXaCT-1 with that from another, unrelated KRAS^G12D+^ sample that had been assayed by the EXaCT-2 assay (Figure S8B). While the EXaCT-1 assay identified this mutation, the confidence in this call was extremely low as there were only 5 total reads covering this position, with only a single read carrying the alternative allele. In comparison, with 772 total reads covering this position, with 86 reads carrying the mutant allele, the EXaCT-2 assay provides far more confidence in both the mutation call and allele frequency (0.11).

##### Comparison of VAF Distributions by Region and Platform

As described in the main text, the distribution of mutational VAFs for the EXaCT-2 assay, along with the TCGA Pancancer project^8^ and MSK-CHORD ^9^ were compared by region type. For example, the “Augmented + Problematic” panel show the VAFs for those mutations which appear in those regions that were augmented in the EXaCT-2 assay and were classified as being problematic by the Genome In a Bottle consortium^10^ along with those regions that were predicted to have a GC content greater than or equal to 80% by the UCSC Genome Browser^11^. Similarly, the “Standard Only” panel displays the VAFs of the mutations that appear in those regions that weren’t predicted to be problematic, and for which the EXaCT-2 uses a standard sequencing coverage as those of other platforms. As such, the sequencing performed by the EXaCT-2 in the two “Standard” sequencing panels is most similar to that which was performed for the TCGA PanCancer Project.

Note, the data for the MSK-CHORD project is completed for completeness, but as shown in figures S9-S11, the distribution of VAFs doesn’t change appreciably between the regions, which is likely due it being a high-depth, targeted panel, as opposed to the EXaCT-2 assay or the platforms used for the TCGA PanCancer project.

As shown in figures S9-S12, the VAFs of the mutations identified by EXaCT-2 assay in the regions of standard coverage display a second mode that is centered near or below 0.1 when the number of reads with supporting the alternative allele (alt reads for short) is less than 15. For example, when the minimum number of alt reads is 5, there is a sizable second mode that is centered near 0.05 (Figure S10). Similarly, when using the slightly more stringent threshold of 10, there still appears to be a small mode, centered near 0.1 (Figure S11). In contrast, the distribution of TCGA VAFs show no such secondary mode, indicating that these secondary modes in the distributions of EXaCT-2 VAFs likely stem from the inclusion of artifactual mutations.

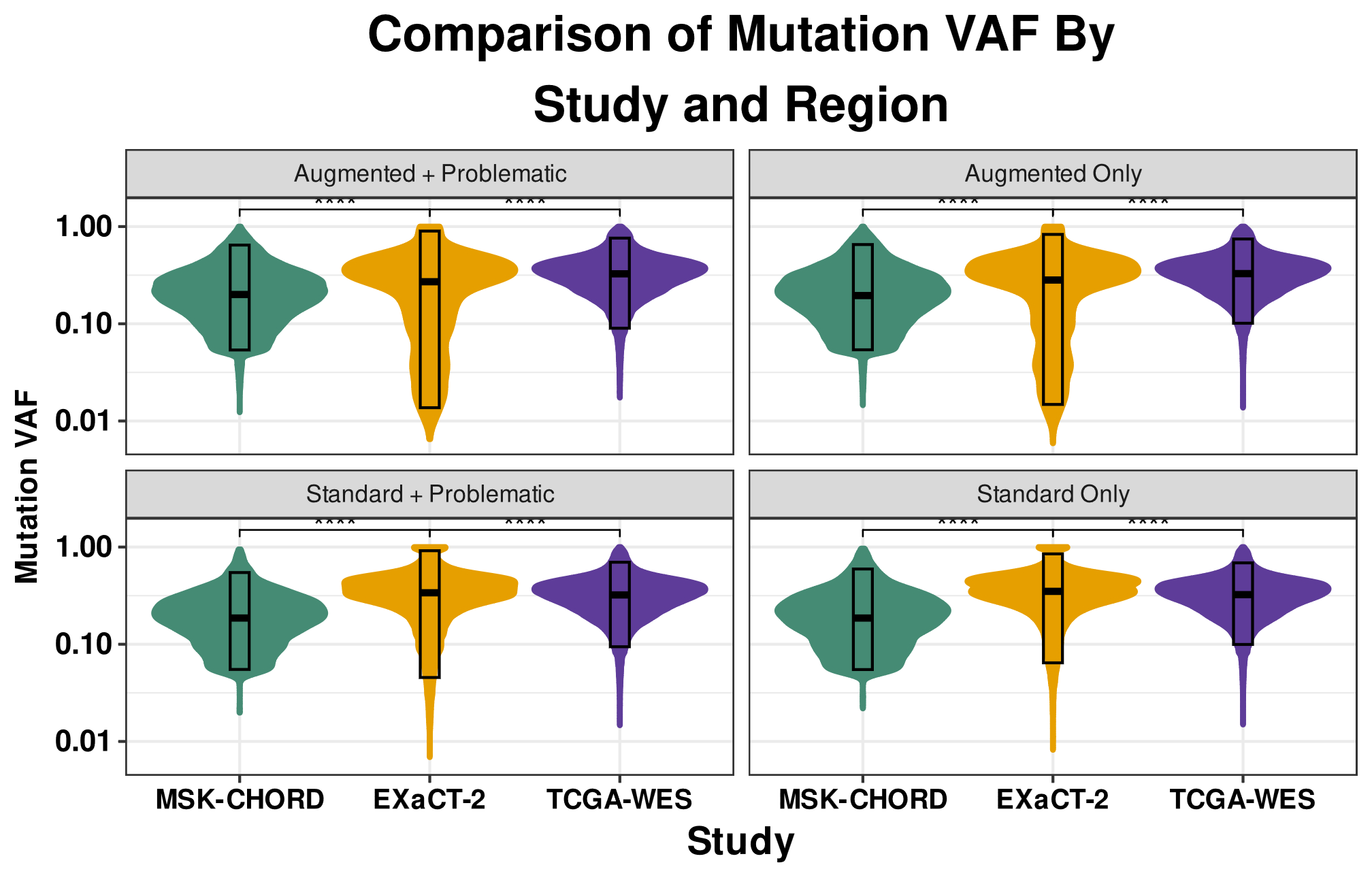

**Figure S9: Distribution of VAFs by Platform for All Mutations Supported by 15 or More Alt Reads.** The distribution of the EXaCT-2 mutational VAFs in the regions of ‘Standard’ coverage closely approximates the distribution of VAFs from the TCGA. In contrast, the distributions of the EXaCT-2 mutational VAFs for the ‘Augmented’ regions demonstrates that the lower tails are clearly lower and broader than is reported by the TCGA, representing the far greater sensitivity offered by the EXaCT-2 platform than standard sequencing assays, even when using a minimum threshold as stringent as requiring 15 alt reads.

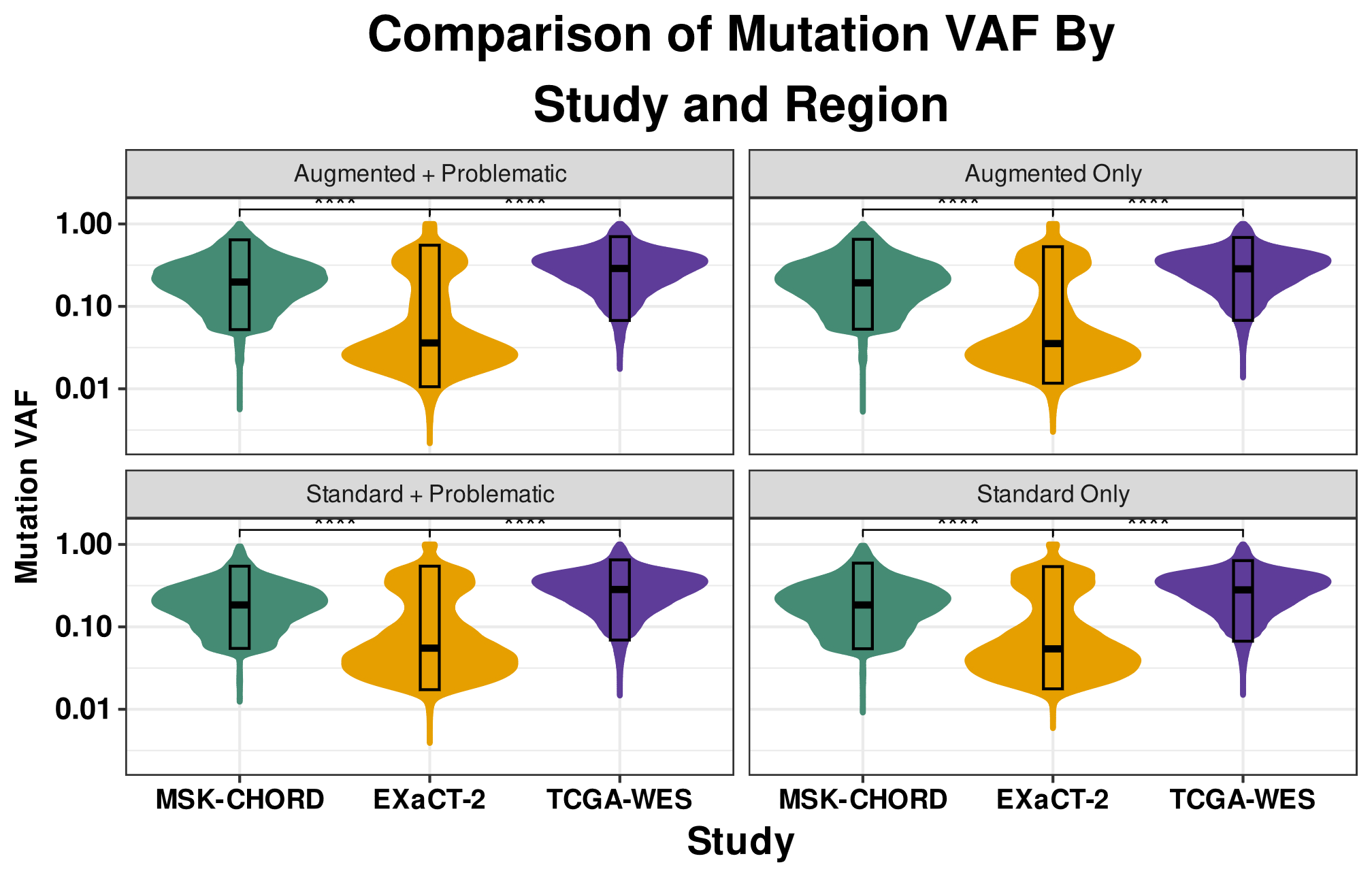

**Figure S10: Distribution of VAFs by Platform for All Mutations Supported by 5 or More Alt Reads.** Unlike the distributions presented in Figure S9, there are secondary, low-VAF modes in the distribution of EXaCT-2 mutational VAFs, regardless of the region, when using a minimum alt read threshold of 5 reads. In contrast, neither the EXaCT-2 nor the TCGA-WES distributions contain these secondary, low-VAF modes. As these secondary modes appear in the regions of standard coverage, this indicates that these distributions include artifactual mutations that are potentially the result of random sequencing errors.

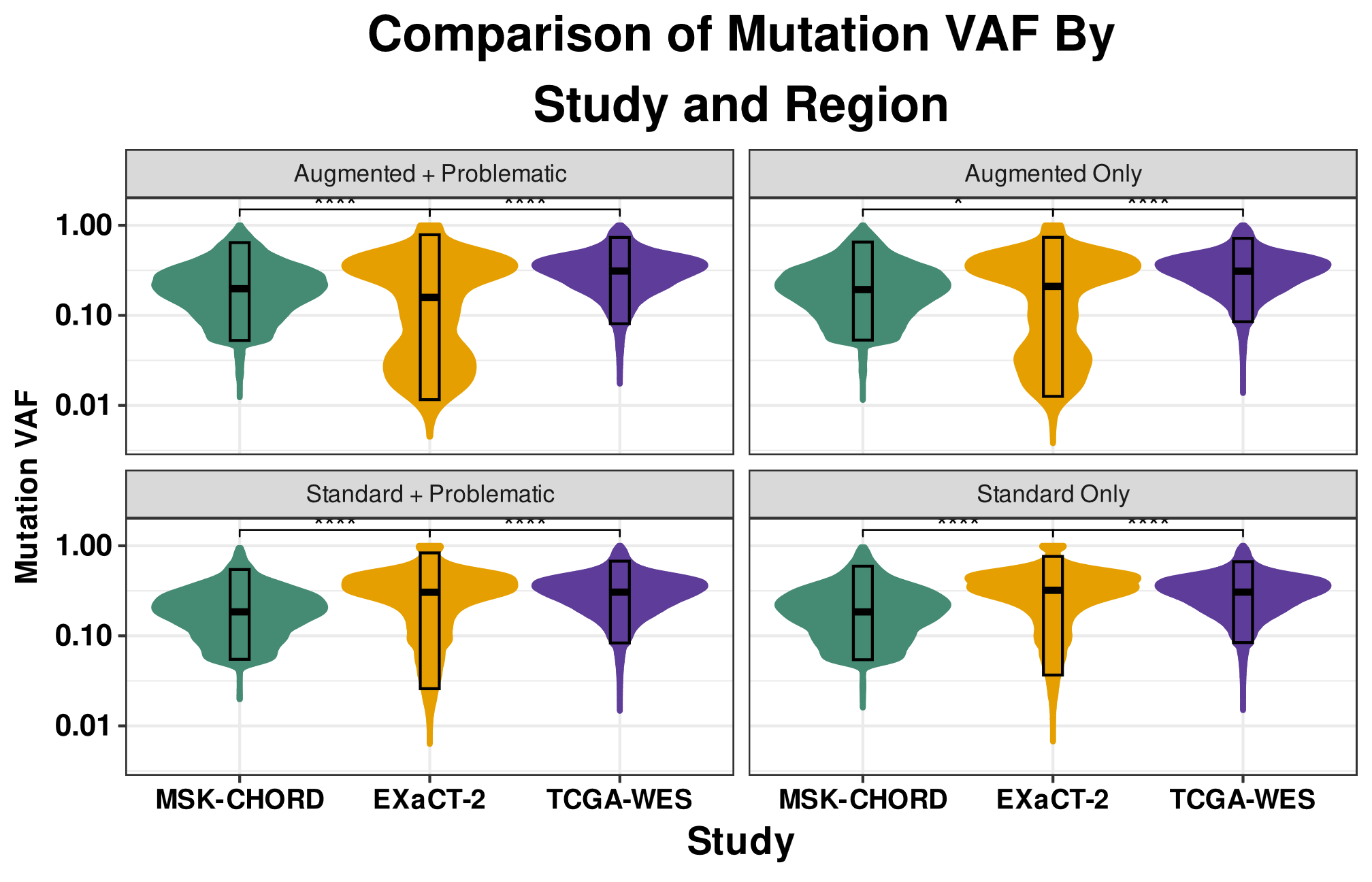

**Figure S11: Distribution of VAFs by Platform for All Mutations Supported by 10 or More Alt Reads.** Unlike the distributions presented in Figure S9, when using a minimum alt read threshold of 10 reads, there are secondary, low-VAF modes in the distribution of EXaCT-2 mutational VAFs in all regions. While the secondary, low-VAF modes in the ‘Standard’ regions aren’t as pronounced as those in Figure S10, they are clearly apparent via visual inspection, indicating that these distributions include artifactual mutations. In contrast, neither the EXaCT-2 nor the TCGA-WES distributions contain these secondary, low-VAF modes.

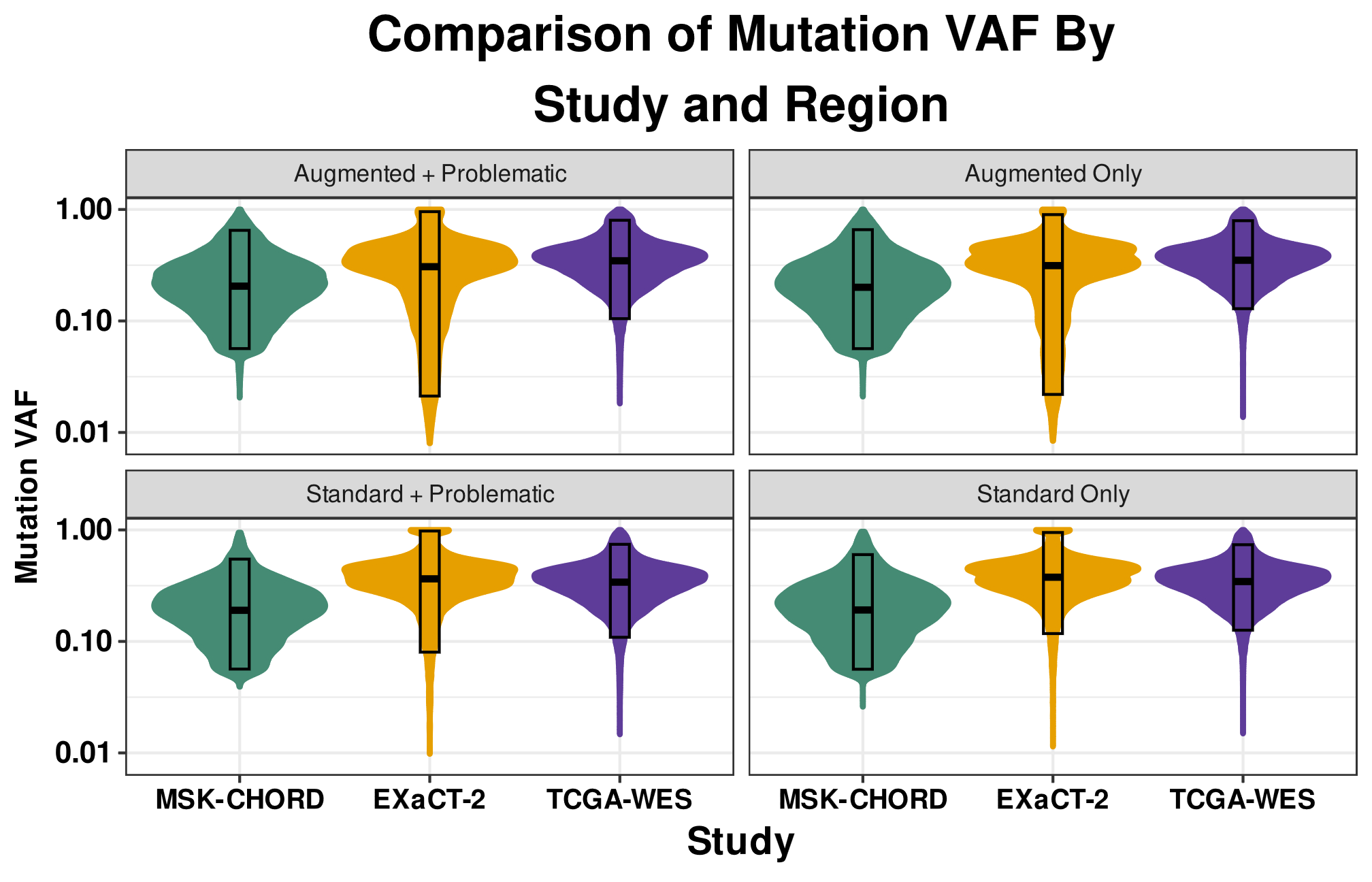

**Figure S12: Distribution of VAFs by Platform for All Mutations Supported by 25 or More Alt Reads.** Included for completeness, this figure demonstrates that further increasing the minimum alt read threshold offers little further benefit to the overall distributions. However, even at this extremely stringent threshold, the broader and deeper distribution of mutational VAFs provided by the EXaCT-2 assay demonstrates that it possesses far greater sensitivity in the augmented regions than is provided by standard sequencing assays.

##### Sensitivity analysis

As an initial test of the EXaCT-2 assay’s accuracy, we performed a sensitivity analysis of germline SNPs and indels using the well-characterized HapMap samples, NA12878 (n=10) and GM24385 (n=1). For this evaluation, the set of SNPs and Indels identified by EXaCT-2 was compared with the expected set of variants for that is provided by the Genome In A Bottle (GIAB) consortium^12^. These evaluations across all 11 samples revealed an average sensitivity of 99.0% and average precision of 99.7%, indicating an exceptionally high concordance and accuracy for the EXaCT-2 and the Gold Standard set that was provided by GIAB (Table S14).

Note, regions that are categorized as FN/FP by GIAB were excluded from this analysis. These regions include highly repetitive regions, or those that are difficult to map, hence we only took into consideration the high confidence regions for our evaluation.

##### Assessment using clinically validated positive controls

As described in the main text, the Weill Cornell Clinical Genomics Lab applied the EXaCT-2 assay to a set of clinically validated samples (n=27) which contained clinically relevant mutations (Figure S13 and Table S15). This test achieved perfect recovery, with a high degree of concordance between the allele frequencies (Pearson correlation: 0.69; p: 5.6e-5). The greatest variation in the observed allele frequencies were associated with the mutations in difficult to assay regions, such as the KRAS^G12+^ and IDH1^R132+^ mutations. As an example, the most discordant mutation we observed had a VAF from the EXaCT-1 assay of 21.4%, while the EXaCT-2 assay estimated the VAF to be over 63%. However, this difference likely stems from the difficulty of standard WES platforms, such as EXaCT-1, to assay this locus, as previously discussed. In this particular case, the EXaCT-1 assay only had access to 14 total reads from which to infer that allele frequency, while EXaCT-2 collected a total of 724 total reads of which 460 carried the alternative allele, thereby providing far more confidence in the estimated allele frequency it generated.

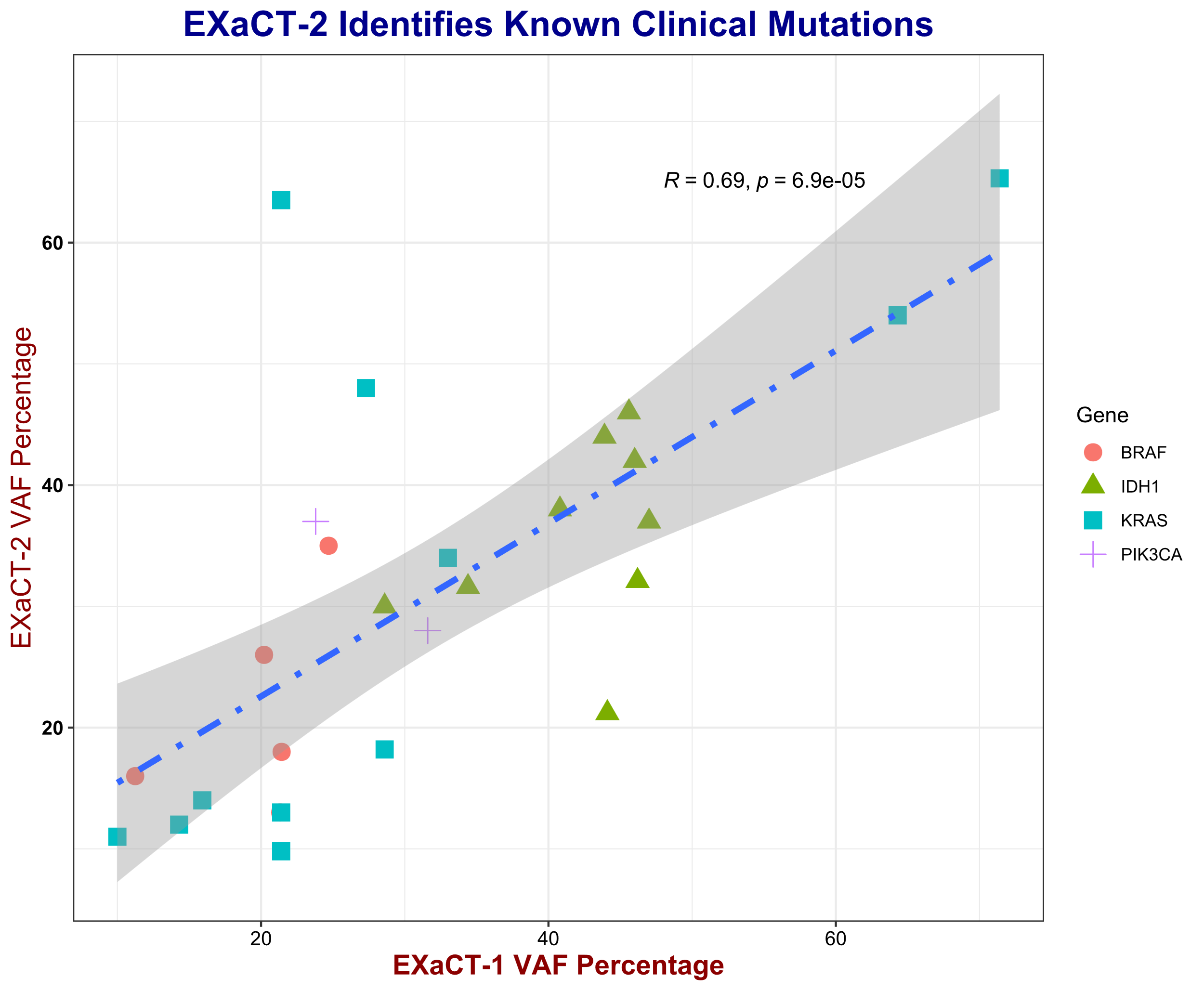

**Figure S13: Comparison of the variant allele frequencies (VAFs) estimated by EXaCT-2 and EXaCT-1.** Distribution of the VAFs estimated by EXaCT-2 and EXaCT-1 for a set of known, experimentally validated and clinically-relevant mutations that were identified in a set of clinical samples. Each point represents a single mutation and the VAF that was estimated by the 2 assays.

##### Customizability of the EXaCT-2 Assay: GTF2I & MTAP Genes

As part of a project to explore the cancer genomes of metastatic Thymoma cancer *(in preparation)*, we customized the EXaCT-2 assay to improve the coverage of 2 well-known driver and marker genes for this cancer ^13,14^. The improvement offered by this customization resulted in significantly improved coverage for both genes (Figure S14) relative to the baseline EXaCT-2 assay, which itself provides a considerably improved coverage than standard whole-exome-based assays such as EXaCT-1.

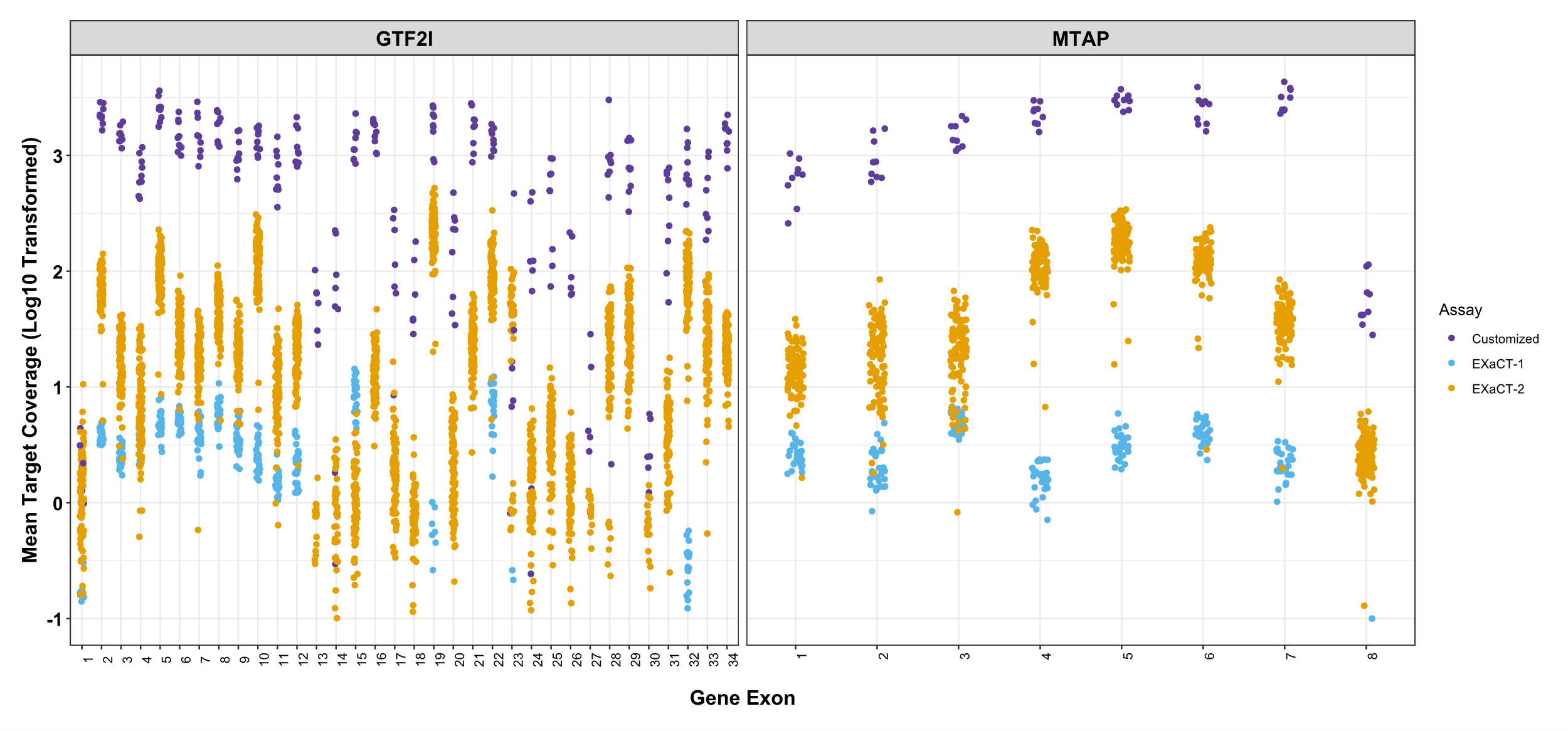

**Figure S14: Enhanced Customization of GTF2I and MTAP Coverage.** This figure presents exon-level coverage for GTF2I and MTAP in samples analyzed using EXaCT-1, EXaCT-2, and a modified EXaCT-2 protocol designed to increase coverage for these genes. The standard EXaCT-2 protocol was followed, with modifications in hybridization using 5µl of EXaCT-2 Target Enrichment Human All Exon biotinylated probes, supplemented with 1µl of a custom MTAP and GTF2I probe cocktail. The customized EXaCT-2 protocol significantly improved coverage compared to both the standard EXaCT-2 and EXaCT-1 assays, which in turn outperform conventional WES methods.

##### TMB in EXaCT-2 dataset compared with the TCGA

As an initial evaluation of the capacity of EXaCT-2 to evaluate Tumor Mutational Burden (TMB) and Microsatellite Instability (MSI), we compared the TMB between the TCGA and EXaCT-2 cohorts, stratifying by tumor of origin. For this comparison (Figure S15), the Tumor Mutational Burden (TMB) was calculated using a total of 106 EXaCT-2 samples and 121 TCGA samples, and to ensure for a meaningful comparison, we limited these cohorts to only include primary tumors. The EXaCT-2 cohort was comprised of 30 samples from Breast cancer, 21 from Diffuse Large B-Cell lymphoma, 6 from Pancreatic Adenocarcinoma, 31 from Lung Adenocarcinoma, and 18 from Bladder cancer. Similarly, the TCGA dataset was comprised of 121 cases of breast cancer, 35 cases of bladder cancer, 13 cases of pancreatic adenocarcinoma, 335 cases of lung adenocarcinoma, and 49 cases of diffuse large B-cell lymphoma. All samples originated from primary tumor sites.^15^

This comparison (Figure S15) indicated that Diffuse Large B-Cell Lymphoma samples exhibit nearly identical average TMB values between the 2 cohorts, while the EXaCT-2 bladder cancer samples exhibited a slightly lower, but statistically similar average TMB to the TCGA bladder samples. In contrast, the EXaCT-2 Lung Adenocarcinoma (LUAD) cohort had a statistically lower average TMB than the TCGA LUAD samples. However, for both the breast (BRCA) and pancreatic adenocarcinoma (PRAD) samples, the EXaCT-2 cohorts had statistically higher average TMB than the TCGA cohorts. While this may appear concerning, superficially, readers should consider that these comparisons are highly imperfect given the different mutation curation and filtering schemes that were employed by the two projects, not to mention the critical differences in the assays that they employed. For example, as previously discussed, a critical feature the EXaCT-2 assay’s design is improved coverage difficult-to-assay regions that were identified by previous projects, such as the TCGA. Furthermore, the TCGA cohort was limited to primary samples, while the EXaCT-2 cohort included primaries as well as organoid and recurrent samples as well.

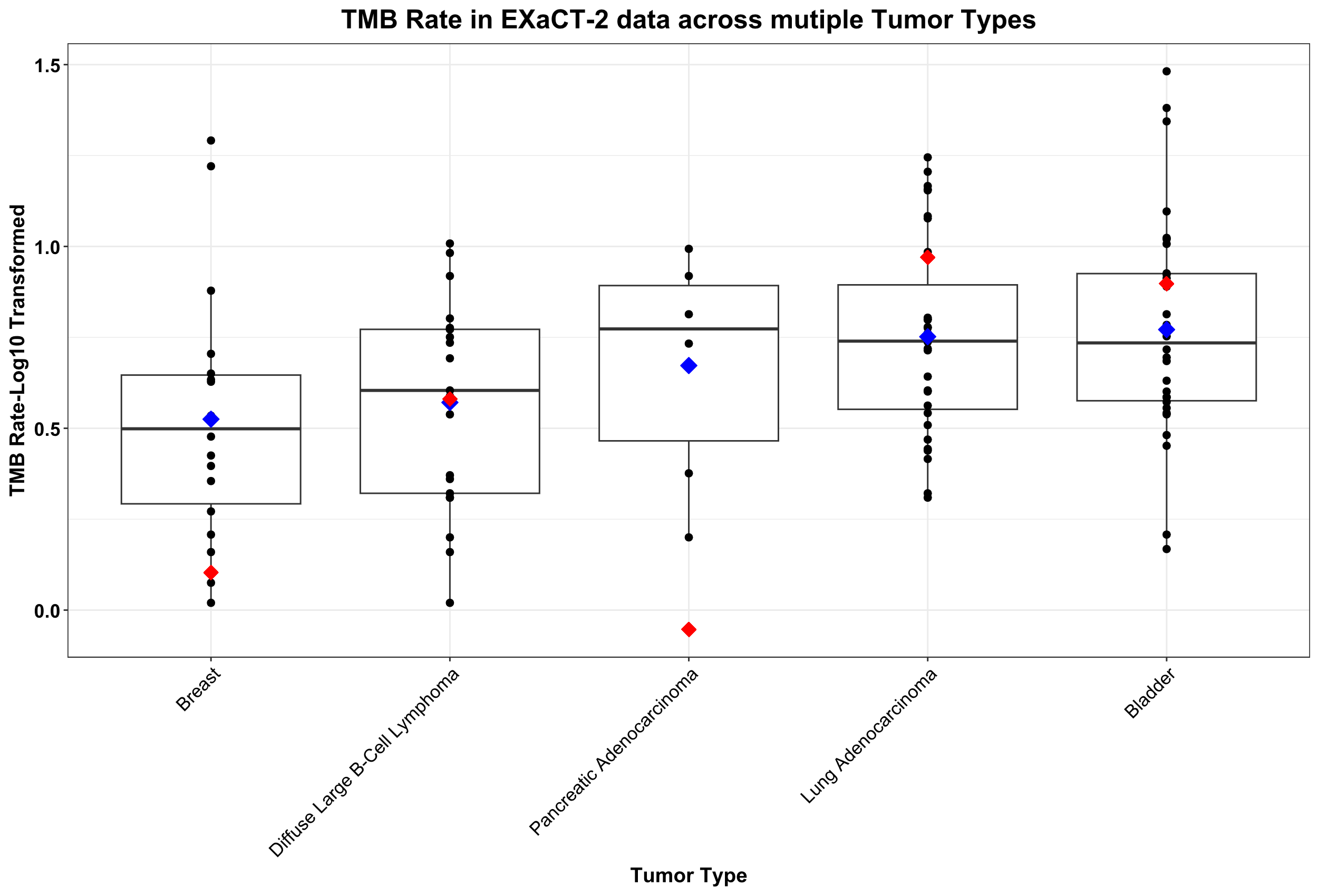

**Figure S15 Comparing TMB from EXaCT-2 dataset to TCGA samples. Each** black dot represents the TMB of an individual sample from EXaCT-2. The blue dot denotes the mean TMB across all samples within each respective tumor type from the EXaCT-2 cohort, while the red dot signifies the mean TMB across all samples from the TCGA cohort for each respective tumor type.

##### TMB vs MSI in EXaCT-2 dataset

In a subsequent evaluation, we calculated the microsatellite instability (MSI) value for the 106 EXaCT-2 samples used for the TMB comparison, using MSIsensor-Pro^16^. As has been observed previously ^17,18^, we observed that samples having a high MSI estimate will likely also have a correspondingly higher TMB estimate; however, in contrast, high TMB status isn’t associated with high MSI status. In these high-TMB, low-MSI cases, it has been speculated that other mechanisms drive the tumor mutational burden ^18,19^.

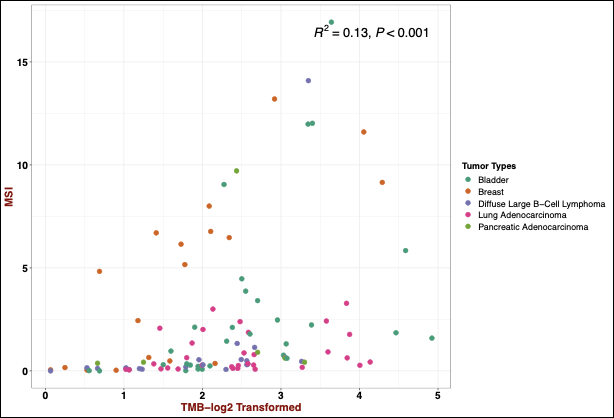

**Figure S16: TMB vs. MSI.** The MSI values for the 106 samples used in Figure S15 are plotted versus the calculated TMB estimates for these samples. As has been shown elsewhere ^17,18^, high MSI values typically correspond to high TMB estimates, while the opposite isn’t true, resulting in a weak, but still significant correlation (Pearson’s R = 0.13, p-value < 0.001).

#### Supplemental Methods:

##### Read alignment and preprocessing

Adapter sequences were removed using fastp, and the trimmed sequencing reads were aligned to the human reference genome (human_g1k_b37) using the Burrows–Wheeler Alignment tool (bwa v0.7.17, using ‘mem’ option). After alignment, PCR duplicates were marked using Sambamba (v0.6.7), followed by realignment around the indels, fixing mates and base quality score recalibration using Genome Analysis Toolkit (GATK, v4.2.0.0), and all unmapped reads were removed (all read with MAPQ equal to 0).

##### Somatic Variant Calling

Somatic variants were identified using a consensus mutation calling approach which integrated seven different mutation callers, this approach is a modified version of the previously published pipeline by the TCGA’s Multi-Center Mutation Calling in Multiple Cancers (MC3) working group ^20^. Our pipeline is comprised of 6 mutation callers (VarScan, SomaticSniper, MuTect2, Strelka2, RADIA, and MuSE) to identify single nucleotide variants (SNVs) and 4 mutation callers (VarScan, MuTect2, Strelka2, Pindel) to identify insertion/deletions (indels). Each caller was run individually on paired tumor and matched normal samples, with the caller-specific set of variants filtered using its own caller-specific criteria, using default settings.

These caller-specific sets of variants were merged into a single vcf file and read depths reported for the same variant called by different tools was averaged. This merged vcf included only those variants that passed the respective caller-specific filtering criteria, and variants were restricted to the genomic coordinates of the capture kit, plus flanking regions equal to 215bp. Note, when averaging the read depts of variants that were called by 2 or more callers, if a variant was called by a given method, but didn’t pass a its internal filtering criteria, the associated read-depth information was excluded from the average.

Finally, this merged set of variants was filtered further based on the following criteria: within-sample allele strand bias, frequency (0.1), total read depth in the tumor (30 total reads, including those with and without the variant), and population frequency (0.01 in either TOPmed (Trans-Omics for Precision Medicine ^21^) gnomAD (Genome Aggregation Database ^22^). Frequently mutated, but biologically unrelated genes such as TTN and the MUC genes were also excluded from the final result (Table S17).

##### Somatic Copy Number Alterations

Somatic Copy Number Alterations (SCNAs) are structural modifications occurring in specific genomic areas, resulting in alterations to the copy number of those regions. These changes are interpreted as either the duplication or deletion of genomic segments. Based on the algorithm from CNVkit ^23^, each genomic segment is assigned a Log2 Ratio, which is obtained by taking the base-2 logarithm of the ratio between the tumor sample's intensity and the matching normal sample's intensity. To categorize these genomic segments as Copy number Gain or Loss, and Amplified or Deleted, we set thresholds on the Log2 ratio as:

- Amplification : >= 1
- Gain : [0.5,1)
- Loss : (-1,-0.5]
- Deletion : <= -1.

Segments with Log2 Ratio < 0.5 or > -0.5, are considered Neutral.

##### Comparing SCNA between EXaCT-1 and EXaCT-2

To assess the SCNA between EXaCT-1 and EXaCT-2, a set of four matching samples from the two assays was selected and compared based on various factors such as Total Fraction Genome Altered (FGA), the total number of segments, and the size of the segments.

FGA: To determine the Total Fraction of genome altered (FGA) in a sample, the sum of the total number of bases with Copy No. Loss or Gain is divided by the total number of bases targeted by EXaCT-2.

Total Number of Segments: When counting the total number of segments, all segments regardless of their copy number status from EXaCT-1 were considered, which were identified using the DNACopy, as previously described ^24,25^. Similarly, the total number of segments from EXaCT-2 encompassed all segments, irrespective of copy number status, obtained from tool CNVKit.

Size of segments: To evaluate the segment size and resolution of copy number segments between EXaCT-1 and EXaCT-2, the segments were categorized into three groups: small segments (<25Mb), mid-sized segments (>25MB and <100Mb), and large segments (>100Mb). The distribution of segments across these three categories was then analyzed.

##### SCNA Gene Concordance

To evaluate the concordance of SCNA and assess the similarity between EXaCT-1 and EXaCT-2 assays, we calculated Gene-Level Concordance for the four corresponding samples. The log2 ratio across each of the 20,000 genes was compared between the matched samples from EXaCT-1 and EXaCT-2. Subsequently, the absolute difference in log2 ratio for each gene between EXaCT-1 and EXaCT-2 matching samples was determined. To mitigate the impact of noisy SCNA calls, a smoothing factor of 0.3 was applied to the absolute difference in log2 ratio. If the absolute difference was less than or equal to 0.3, an average of the log2 ratio for that gene from EXaCT-1 and EXaCT-2 was computed. If the absolute difference exceeded 0.3, the original log2 values for the genes were retained. Following this, genes were categorized into different types of SCNA Events (Copy No. Gain or Copy No. Loss or Neutral) based on a predefined threshold.

Detection of Clonal Rearrangements

To identify all clonal rearrangements within a sample, the MiXCR tool (version 4.6.0) ^26^ was employed. The dataset consisted of whole-exome paired-end samples, and the following command line was utilized to extract clonotype information:

*mixcr analyze exome-seq --species hsa --no-reports --no-json-reports Fastq1 Fastq2 sampleID*

*mixcr exportClones --dont-split-files sampleID.contigs.clns sampleID.tsv*

In the above command, 'Fastq1' and 'Fastq2' contain reads from the forward and reverse strands, respectively, of the DNA fragments. 'sampleID' represents the sample name used for the output files. The generated output files include 'sampleID.contigs.clns' and 'sampleID.tsv', with the latter containing read fraction, nucleotide, and amino acid clonotype sequences, as well as Phred quality information for each clonotype present in the sample.

The predominant clonotype in the sample is typically identified based on the highest read fraction, indicating its abundance.Further analysis involves computing the Read frequency, calculated as *read frequency = read fraction * 100*, which represents the percentage of all reads in the sample assigned to that specific clonotype.

##### Depth of Coverage

EXaCT-2 has been specifically designed to achieve increased coverage in select genomic regions crucial for cancer studies. To assess and highlight the increased coverage in these Augmented regions compared to Non-Augmented regions, a set of germline samples were analyzed across these two sets of regions. The comparison involved computing the Mean Target Coverage in both sets of regions using Picard CollectHsMetrics tool ^27^, where Mean Target Coverage represents the average depth across a specific set of targeted regions. Here, two sets of regions include: Augmented regions, which encompass clinically relevant areas designed to have enhanced coverage in A, including intronic and intergenic regions. The other set comprises Non-Augmented regions, covering the remaining areas targeted by EXaCT-2.

The comparison of mean target coverage in both sets of regions was conducted and categorized based on DIN quality and Total Reads present in the sample to explore the relationship between mean target coverage, total reads, and DIN across different genomic regions targeted by EXaCT-2.

###### Categorized based on Total On-Target Reads

To further emphasize and highlight into the depth of coverage for EXaCT-2, and underscore distinctions between Augmented and Non-Augmented regions on the same set of germline samples as previously analyzed, the mean target coverage in these two sets of regions was classified based on an additional criterion: Total On-Target Reads, where Total On-Target Reads encompass only those reads that fall within the specified target regions. The input BAM file is filtered to include only regions falling under Augmented and Non-Augmented categories targeted by EXaCT-2. Subsequently, Picard is used to compute the total on-target reads for both Augmented and Non-Augmented regions, excluding reads that do not align to these specific regions.

Quantiles of the total on-target reads were then determined as follows: samples with reads falling <= 25% (<=102.7 M reads); samples with reads falling > 25% and <= 50% (>102.7 M and <=123 M reads); samples with reads falling > 50% and <= 75% (>123 M and <=141.5 M reads); and samples with reads falling > 75% (>141.5 M reads).

###### Enhanced coverage in clinically Relevant Genes

EXaCT-2 has also been designed to enhance coverage in clinically relevant genes. To show that EXaCT-2 has achieved higher coverage in these specific genes compared to EXaCT-1, a subset of genes of interest was considered and compared between the two assays. Mean Target Coverage was calculated for each gene, and the coverage was compared across the same set of germline samples as previously analyzed.

###### Comparison of mean target coverage for KRAS G12* mutations

We downloaded the read coverage for KRAS G12* mutations TCGA (WES and WGS) and MSK-IMPACT cohorts from cBioPortal. In total, there were 2,671 KRAS positive samples from cBioPortal and we compared with the coverage of 61 KRAS positive EXaCT-2 samples.

##### Customization for complex genomic regions

###### Regions with high GC content

EXaCT-2 is also purposefully designed and can be tailored to achieve enhanced coverage in challenging genes that pose difficulties for various reasons. One of the potential problematic regions includes those with high GC content, such as in the case of the CEBPA gene. EXaCT-2 addresses this challenge by incorporating additional probes to cover these regions with significantly higher coverage compared to EXaCT-1. To demonstrate and substantiate this claim, we investigated the same region with high GC content (>= 80%) in the Integrative Genomics Viewer (IGV) ^28^ and compared the read counts in a sample from EXaCT-2 with a sample from EXaCT-1 within that region.

##### Total Mutation Burden (TMB) estimation

The calculation of TMB involves dividing the total count of mutation events by the number of bases per million in the specified coding regions of EXaCT-2. This process specifically considers certain SNVs and Indels, following predefined filters. Only mutations falling into variant classes, including Frameshift Insertions/Deletions, Inframe Insertions/Deletions, Start and Stop codon Mutations, Splice-Site Mutations, Missense mutations, Splice mutations, Non-Coding, and Synonymous Mutations, are considered.

Moreover, these mutations are subjected to additional filters, requiring Tumor Total Depth >= 30, Control Total Depth >= 30 and Tumor VAF >= 0.1. Furthermore, Genome Aggregation Database (gnomAD) Allele Frequency <= 0.01, and Trans-Omics for Precision Medicine (topmed) Allele Frequency <=0.01, ensuring the exclusive inclusion of rare variants. Additionally, variants must have proper HGVSP and HGVSC notations. Genes and variants identified as artifacts, based on a predefined list, are also excluded from the analysis.

##### Microsatellite Instability (MSI) Detection

MSI serves as a predictive and prognostic biomarker indicating DNA mismatch repair deficiency, a condition frequently observed in cancer. Here, MSISensor-Pro ^16,29^ is used which utilizes the length distributions of microsatellites at specific loci in both the tumor and the matched-normal from the BAM files followed by statistical comparisons on the observed distributions in both the samples.

Using the tool, it identifies the total number of sites with sufficient data (a minimum of 20 reads spanning both tumor and normal samples) and then determines the number of somatic sites. The MSI score is then computed, representing the percentage of somatic sites. According to prior literature, samples with an MSI score of >= 0.4 are classified as MSI-high.
